# Exploring pleiotropy in Mendelian randomisation analyses: What are genetic variants associated with “cigarette smoking initiation” really capturing?

**DOI:** 10.1101/2023.08.04.23293638

**Authors:** Zoe E. Reed, Robyn E. Wootton, Jasmine N. Khouja, Tom G. Richardson, Eleanor Sanderson, George Davey Smith, Marcus R Munafò

## Abstract

**Background:** Genetic variants used as instruments for exposures in Mendelian randomisation (MR) analyses may also have horizontal pleiotropic effects (i.e., influence outcomes via pathways other than through the exposure), which can undermine the validity of results. We examined the extent to which horizontal pleiotropy may be present, using smoking behaviours as an example.

**Methods:** We first ran a phenome-wide association study in UK Biobank, using a genetic instrument for smoking initiation. From the most strongly associated phenotypes, we selected those that we considered could either plausibly or not plausibly be caused by smoking. We next examined the association between genetic instruments for smoking initiation, smoking heaviness and lifetime smoking and these phenotypes in both UK Biobank and the Avon Longitudinal Study of Parents and Children (ALSPAC). We conducted negative control analyses among never smokers, including children in ALSPAC.

**Results:** We found evidence that smoking-related genetic instruments (mainly for smoking initiation and lifetime smoking) were associated with phenotypes not plausibly caused by smoking in UK Biobank and (to a lesser extent) ALSPAC, although this may reflect the much smaller sample size in ALSPAC. We also observed associations with several phenotypes among never smokers.

**Conclusion:** Our results suggest that genetic instruments for smoking-related phenotypes demonstrate horizontal pleiotropy. When using genetic variants – particularly those for complex behavioural exposures – in genetically-informed causal inference analyses (e.g., MR) it is important to include negative control outcomes where possible, and other triangulation approaches, to avoid arriving at incorrect conclusions.

## Introduction

Mendelian randomisation (MR) is a genetically-informed causal inference approach that uses SNPs identified in GWAS of putative exposures to examine causal effects of those exposures on outcomes [1, 2]. A key assumption for MR is that these single nucleotide polymorphisms (SNPs) are associated only with the outcome through the exposure of interest, and not via other pathways. If this assumption is not met then this may indicate the presence of pleiotropy, which may invalidate MR analyses, meaning that resulting causal inferences could be erroneous [3–5].

Pleiotropy can take two different forms: *vertical* pleiotropy and *horizontal* pleiotropy, with the latter including *correlated horizontal* pleiotropy. These are shown in Fig 1. Vertical pleiotropy (Fig 1a), whereby the genetic variants operate via an intermediate phenotype, is not problematic for MR. Horizontal pleiotropy, however, undermines a key assumption of MR [6]. Horizontal pleiotropy (Fig 1b) can be balanced (i.e., the pleiotropic effects have a net effect of zero due to SNPs acting through different pathways resulting in both positive and negative effects that negate each other) or unbalanced (i.e., where the net effect is not zero). Balanced horizontal pleiotropy will increase the heterogeneity in MR analyses, but does *not* bias the effect estimates obtained; however, unbalanced horizontal pleiotropy is problematic and biases the results obtained. If such unbalanced pleiotropic pathways exist, then the results of MR studies may not be valid. Horizontal pleiotropy may also be *correlated* (Fig 1c). In this case, the genetic instrument influences a heritable confounder that influences both the exposure and outcome. In an extreme case, the SNPs may act entirely via a different primary phenotype, meaning that the target phenotype has been mis-specified (Fig 1d).

**Fig 1.**
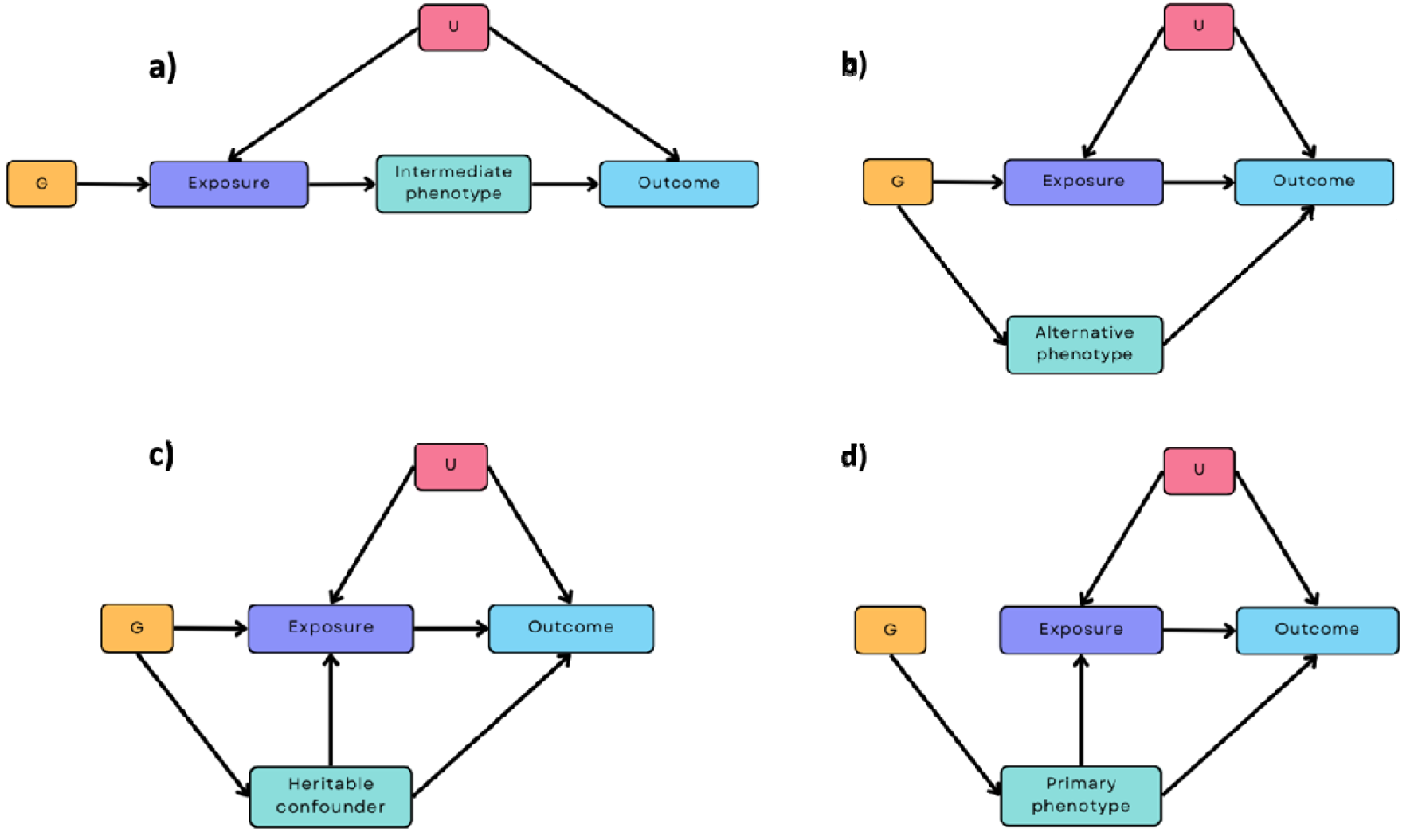
Different pleiotropic pathways to consider when estimating the causal effect of an exposure on an outcome using Mendelian randomisation (MR).

Panel a) shows a vertical pleiotropic pathway where the genetic instrument G is associated with the outcome via an intermediate phenotype. Panel b) shows a horizontal pleiotropic pathway, where G is associated with both the exposure and another phenotype on a different pathway. Panel c) shows a correlated horizontal pleiotropic pathway, where G is also associated with a heritable confounder that, in turn, influences the exposure as well as the outcome [7]. Panel d) shows a pathway where the exposure is mis-specified, where G is related to a heritable confounder only, but this causes both the exposure and the outcome. Scenario a) does not invalidate MR estimates because the intermediate phenotype is on the causal path from the exposure to the outcome and the primary phenotype (exposure) is specified correctly [8], whereas scenarios b) to d) may invalidate the results of MR analyses. G = genetic instrument; U = heritable confounders of exposure-outcome relationship

Such horizontal pleiotropic effects may occur as a result of the much larger samples used in genome-wide association studies (GWAS) in recent years, identifying weaker associations with SNPs than found in smaller samples and increasing the likelihood that SNPs included in genetic scores are predictive of broader, correlated phenotypes [9]. As noted above, this is problematic for MR if the SNPs we use to instrument an exposure are in fact influencing other phenotypes via horizontal pleiotropy. Therefore, it is important to ascertain whether the SNPs we use as genetic instruments for an exposure of interest are in fact instrumenting only this exposure, or whether there is evidence of horizontal pleiotropy. Whilst several MR sensitivity methods now exist that can help us to infer the likelihood of bias from horizontal pleiotropy and identify whether correlated pleiotropy is present [7, 10, 11], it is difficult to directly test for pleiotropic effects without understanding more about the functional biological effects of these SNPs.

One example of a complex behavioural exposure where horizontal pleiotropy may be operating is cigarette smoking. Recent GWAS have identified many genetic variants associated with different smoking behaviours (e.g., smoking initiation, heaviness of smoking). However, subsequent studies have suggested that some of these SNPs may also be influencing different phenotypes via independent pathways (i.e., not via smoking). For example, in studies by Khouja and colleagues [12] and by Schellhas and colleagues [13], SNPs for smoking initiation were also found to be associated with risk-taking behaviours in young adults, personality traits in adults and adolescents, and externalising disorders in children at age 7.

We assessed the extent that horizontal pleiotropy may be operating, using smoking behaviours as an example. Specifically, we investigated whether phenotypes that we considered could either plausibly be caused by smoking, or could not plausibly be caused by smoking, were associated with genetic risk scores for smoking initiation, smoking heaviness and a lifetime smoking index. We examined this across two cohorts, UK Biobank and the Avon Longitudinal Study of Parents and Children (ALSPAC), which are subject to different patterns of selection bias. We also conducted negative control analyses in never smokers, where any genotype-outcome associations could not be mediated via smoking [14, 15].

Associations with outcomes that could not plausibly be caused by smoking may provide evidence: 1) of horizontal pleiotropy (either correlated or uncorrelated), or 2) that our exposure phenotypes are mis-specified (i.e., that the exposure is not actually the primary phenotype and the the genetic variants used as the instrument are associated with, for example, a downstream phenotype).

## Methods

Our study consists of two parts, a discovery phase (a phenome-wide association study in UK Biobank), and an analysis phase (polygenic risk score analyses in UK Biobank and ALSPAC).

### Pre-registration

We pre-registered our analysis plan on the Open Science Framework (https://doi.org/10.17605/OSF.IO/37XYN). We stated that we would examine evidence of pleiotropy in two follow-up studies using the Million Veteran Program (MVP) and the Avon Longitudinal Study of Parents and Children (ALSPAC). However, in a deviation from our pre-registered analyses, we only conducted analyses in ALSPAC due to data availability, and because we could also use child data in ALSPAC prior to smoking commencing as negative control analyses. Additionally, we did not conduct analyses using different p-value thresholds, as stated in our pre-registration, as we decided it was more relevant to see whether genome-wide significant SNPs specifically (i.e., p<5×10^-08^), often used as instruments in approaches such as MR, show evidence of horizontal pleiotropy.

### Study cohorts

#### UK Biobank

UK Biobank is a large population-based prospective health research resource with around 500,000 participants, aged 38 to 73 years at recruitment (between 2006 and 2010), from across the UK [16]. A range of data have been collected including sociodemographic data, lifestyle, cognitive function, self-reported measures and physical and mental health measures, with the aim of identifying determinants of human disease. Data have been collected via several methods, including paper- and web-based questionnaires, computer assisted interviews, clinic visits and data linkage. Baseline assessment took place across 22 assessment centres to enable recruitment from a range of locations, but further data collection is ongoing. Further information can be found on the UK Biobank website (www.ukbiobank.ac.uk). UK Biobank received ethics approval from the Research Ethics Committee (REC reference for UK Biobank is 11/NW/0382). We excluded participants who withdrew their consent using the latest withdrawal lists for this project (project number: 16729). We restricted analyses to individuals who self-reported as ‘White’ and ‘British’ and who had very similar genetic ancestry based on a principal components analysis of genotypes, which aims to minimise variation in non-genetic and genetic factors. The self-reported responses were from questions in the touchscreen questionnaire asking, ‘What is your ethnic group?’ with the options of White; Mixed; Asian or Asian British; Black or Black British, Chinese, Other ethnic group, Do not know; Prefer not to answer. If they selected ‘White’ then they were asked ‘What is your ethnic background?’ with the options of British; Irish; Any other white background; Prefer not to answer. We note that ethnicity is a complex social construct that can have different meanings across different contexts, and is distinct to, although often overlapping with, genetic ancestry [17]. We also removed related individuals or those with mismatched sex.

#### ALSPAC

Pregnant women resident in Avon, UK with expected dates of delivery 1^st^ April 1991 to 31^st^ December 1992 were invited to take part in the study. The initial number of pregnancies enrolled was 14,541. Of these, there was a total of 13,988 children alive at age 1. When children were age 7, additional eligible cases who had failed to join the study originally were recruited, resulting in a total sample size of 14,901 children. There are 14,833 mothers enrolled in ALSPAC and 3,807 partners [18]. ALSPAC is described in more detail in the cohort profile papers [19, 20]. The study website contains details of the available data through a data dictionary and variable search tool (http://www.bristol.ac.uk/alspac/researchers/our-data/). We used data from children, mothers and fathers/partners (not all were male) in our analyses. Ethics approval was obtained from the ALSPAC Ethics and Law Committee and the Local Research Ethics Committees (http://www.bristol.ac.uk/alspac/researchers/research-ethics/). Consent for biological samples was collected in accordance with the Human Tissue Act (2004). Informed consent for the use of data collected via questionnaires and clinics was obtained from participants following the recommendations of the ALSPAC Ethics and Law Committee at the time.

### Phenotypic measures

#### Smoking related exposures

We used three smoking related exposures in our analyses to examine the extent of pleiotropy across these: smoking initiation (i.e., ever versus never smoked), smoking heaviness (as measured by cigarettes smoked per day), and a lifetime smoking index (only available in UK Biobank). Further details of these measures for UK Biobank and ALSPAC can be found in the Supplementary Materials (Section 1).

#### Discovery phase

We ran a phenome-wide association study (PheWAS) [21] for smoking initiation using a polygenic risk score (PRS) of smoking initiation as the exposure (p<5×10^-08^), constructed in UK Biobank from publicly available GWAS data (excluding UK Biobank) [22]. Further details on the PheWAS can be found in the Supplementary Materials (Section 2). We used smoking initiation only for the PheWAS, as we hypothesised that this behaviour would be most likely to be associated with phenotypes not plausibly caused by smoking. From the most strongly associated phenotypes (i.e., the top 100 results, based on p-values, see Supplementary Table S1), we selected several phenotypes that could plausibly be caused by smoking, and those that could not plausibly be caused by smoking, in subsequent analyses.

Phenotypes were selected on the basis that they were: 1) likely to be*caused* by smoking (e.g., chronic obstructive pulmonary disease, COPD), or 2) that they may be associated with, but were *not likely* to be caused by, smoking (e.g., age of mother at time of questionnaire). These were selected from consensus between authors, based on their expert knowledge and previous studies (see Supplementary Materials Section 3).

#### Phenotypes in UK Biobank plausibly caused by smoking for analysis phase

Details of the 13 phenotypes plausibly caused by smoking are shown in Supplementary Table S2. These were: body mass index (BMI); body fat percentage; wheeze; C-reactive protein (CRP); ever reported COPD; had dentures; overall health rating; gamma glutamyl transferase (GGT); white blood cell count; mean sphered cell volume; seen GP for nerves, anxiety, or depression; numbers of medications taken; and alcohol consumption. We transformed continuous variables which were highly positively skewed to be normally distributed using an inverse normal rank transformation (INRT) (as indicated in Supplementary Table S2).

#### Phenotypes in UK Biobank not plausibly caused by smoking for analysis phase

Details of the 13 phenotypes not plausibly caused by smoking are shown in Supplementary Table S3. These were: lifetime number of sexual partners; younger age at first live birth; Townsend deprivation index; takes part in a religious group; cereal intake; risk-taking; time spent watching television; liking for cabbage; mobile phone usage; ease of skin tanning; mother’s age at time of questionnaire; back pain and had an operation on the left-side of the body. Variables we transformed using INRT are indicated in Supplementary Table S3.

#### Potential confounders in UK Biobank for analysis phase

Potential confounders of our genetic instrument and outcomes were: age at assessment centre attendance, self-reported sex and the first 10 principal components (PCs) of population structure from genotype data.

#### Phenotypes in ALSPAC plausibly and not plausibly caused by smoking for analysis phase

Where the phenotypes plausibly or not plausibly caused by smoking (or sufficiently similar phenotypes) were available in ALSPAC, we used these to examine whether UK Biobank results could be due to selection biases specific to UK Biobank, or whether these associations are observed in other cohorts. We used data from mothers, fathers/partners, and children at age 10, the latter to examine whether associations existed prior to smoking onset, as this would provide evidence that pathways are not via the exposure of own smoking.

Details on the phenotypes used can be found in Supplementary Tables S4 and S5. The measures used, and time points are summarised in Fig 2. We generally used the earliest time point (T1) where data was available. However, there were some cases where this was less appropriate (e.g., due to pregnancy or early child age), in which case we ran additional analyses with the next time point (T2) to assess consistency (see Supplementary Tables S4 and S5).

**Fig 2.**
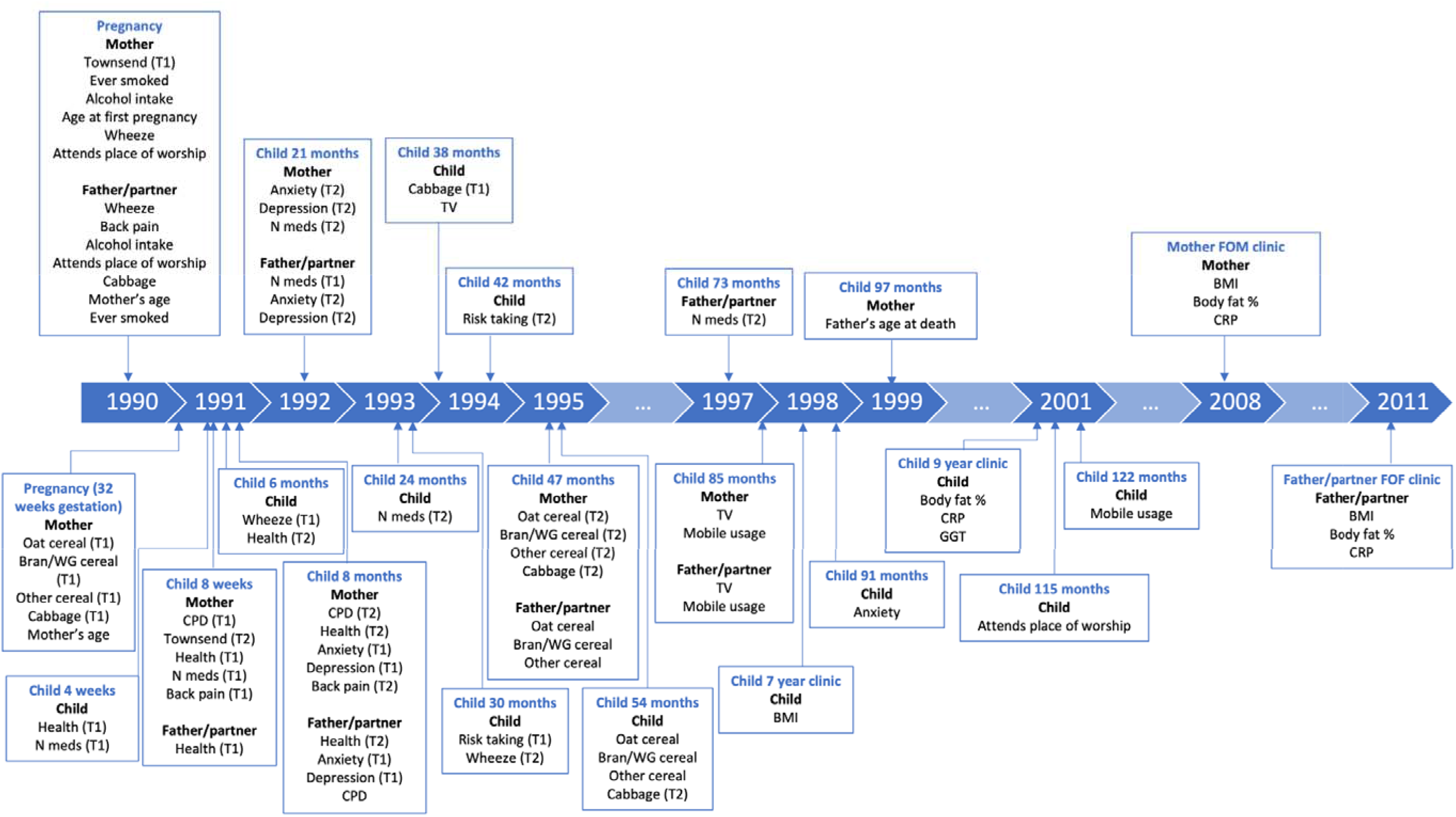
Measures used in ALSPAC.

For continuous phenotypes, we typically removed outliers by excluding individuals from analyses where data was three times the interquartile range above the upper quartile or below the lower quartile. However, we did not remove outliers for number of medications as values all seemed plausible.

### Genetic data for analysis phase

#### UK Biobank

Of the 488,377 participants with genotyped samples, 336,988 were included in analyses (quality control steps are described in the Supplementary Materials, Section 4).

#### ALSPAC

Genetic data were obtained from a combination of blood and buccal samples (see Supplementary Materials Section 5). After quality control and removing those who had withdrawn consent, there were 7,961 children, 7,912 mothers and 1,722 fathers/partners with genotype data available.

### Polygenic risk score construction for analysis phase

#### UK Biobank

To construct PRS we used a 10-fold cross validation approach to reduce potential overfitting bias due to sample overlap as UK Biobank data was used for the discovery GWAS and PRS construction [23]. This involved randomly splitting the UK Biobank population into 10 subsamples and then running 10 GWAS for each of the smoking related exposures in turn on 90% of the UK Biobank sample (see Supplementary Materials Section 6), with a different 10% of the sample removed each time. PRS were then constructed for this remaining 10% of the sample to avoid sample overlap (see Supplementary Materials Section 7). After 10 iterations were complete, PRS from each of the 10% subsamples were brought together such that cross-validated scores for each of the smoking related exposures were available in the full sample. Further details on this 10-fold cross validation methodology and related simulations are reported elsewhere [24].

#### ALSPAC

To construct PRS in ALSPAC, we used SNPs and weights from GWAS of smoking heaviness and initiation we conducted in UK Biobank (without the 10-fold cross validation approach, see Supplementary Materials Section 8) and from a published GWAS of lifetime smoking also conducted in UK Biobank [25]. This allowed us to replicate our UK Biobank analyses.

### Statistical analysis in analysis phase

All analyses were conducted in R version 3.5.1.

#### Polygenic risk score analyses in UK Biobank

We tested the association of PRS for smoking initiation, smoking heaviness, and lifetime smoking index (constructed using our cross-validation approach) with the phenotypes plausibly and not plausibly caused by smoking. We used linear regression models for continuous outcomes, logistic regression models for binary outcomes and ordinal logistic regression for ordered factor outcomes. All models were adjusted for age, sex and the first 10 PCs. We stratified analyses using the smoking heaviness PRS by smoking status (never, former and current smokers), where analyses in never smokers acted as a type of negative control analysis, because individuals have not been exposed to their own smoking.

#### Polygenic risk score analyses in ALSPAC

Similar to UK Biobank analyses, we tested the association of the PRS for smoking initiation, smoking heaviness and lifetime smoking on the phenotypes plausibly and not plausibly caused by smoking available in ALSPAC, separately for mothers, fathers/partners and children (due to phenotypic differences). Here we used GWAS from UK Biobank to replicate our analyses in UK Biobank. All models were adjusted for age, sex (in children only) and the first 10 PCs. We stratified analyses with the smoking heaviness PRS by smoking status in adults (ever or never smokers). Stratifying by smoking status in these analyses could introduce collider bias if a confounder affects both the outcome and smoking status – making smoking status a collider. Therefore, as well as conducting the analyses on adults, we additionally conducted the analyses in children (who will not have started smoking) where collider bias would not be introduced.

### Data availability

GWAS data for smoking initiation with UK Biobank and 23andMe removed can be found here: https://conservancy.umn.edu/handle/11299/201564. Full GWAS summary statistics for the 23andMe discovery data set (which we combined with the publicly available smoking initiation data) will be made available through 23andMe to qualified researchers under an agreement with 23andMe that protects the privacy of the 23andMe participants. Please visit https://research.23andme.com/collaborate/#dataset-access/ for more information and to apply to access the data.

ALSPAC data access is through a system of managed open access (http://www.bristol.ac.uk/alspac/researchers/access/).

UK Biobank data are available through a procedure described at http://www.ukbiobank.ac.uk/using-the-resource/.

### Code availability

Analysis code is available from the University of Bristol’s Research Data Repository (http://data.bris.ac.uk/data/), DOI: To be added on publication

## Results

### UK Biobank sample characteristics

Sample characteristics for participants included in this study (N=101,397 to 336,988) are shown in Supplementary Table S6. The mean age was 57 years (SD=8) and 54% were female. There were 45% participants who had ever smoked, with an average of 5 (SD=10) cigarettes per day and a lifetime smoking score average of 0.34 (SD=0.68). A 1 SD increase in this score is, for example, equivalent to being a current smoker who has smoked 5 cigarettes per day for 12 years, or a former smoker who smoked 5 cigarettes per day for 21 years but stopped smoking 10 years ago, compared to a never smoker.

### Polygenic risk score associations in UK Biobank (analysis phase)

#### Phenotypes plausibly caused by smoking

Results from analyses between each of the three smoking-related PRS and the phenotypes plausibly caused by smoking are shown in Fig 3 and Supplementary Table S7. Results for smoking heaviness are presented for each of the three categories (never, former and current smokers). We found evidence of associations between the PRS for lifetime smoking and PRS for smoking initiation and all of our phenotypes plausibly caused by smoking (p-values=1.06×10^-29^ to 2.77×10^-03^ for lifetime smoking and p-values=1.82×10^-18^ to 4.48×10^-04^ for smoking initiation), with the direction of effect consistent for both PRS. For the smoking heaviness PRS we found associations with some but not all of the phenotypes plausibly caused by smoking. In some cases, the effect estimates were attenuated compared to the other smoking PRS associations; however, confidence intervals were also generally wider for smoking heaviness so we may have had lower power to detect these effects. For former smokers, we found positive associations with the smoking heaviness PRS and CRP (b=0.007; 95% CI: 0.001 to 0.01; p=0.02), COPD (OR=1.04; 95% CI: 1.007 to 1.07; p=0.01) and poorer health (OR=1.01; 95% CI: 1.00 to 1.03; p=0.02) only. For current smokers, we found negative associations with the smoking heaviness PRS and BMI (b=-0.10; 95% CI: −0.15 to −0.05; p=2.19×10^-04^) and body fat percentage (b=-0.15; 95% CI: −0.22 to −0.08; p=4.31×10^-05^). This is the opposite direction to the associations with the lifetime smoking and smoking initiation PRS. We also found a positive association with COPD (OR=1.06; 95% CI: 1.03 to 1.10; p=5.06×10^-04^). For never smokers, we did not find evidence of any associations between the phenotypes plausibly caused by smoking and the smoking heaviness PRS.

**Fig 3.**
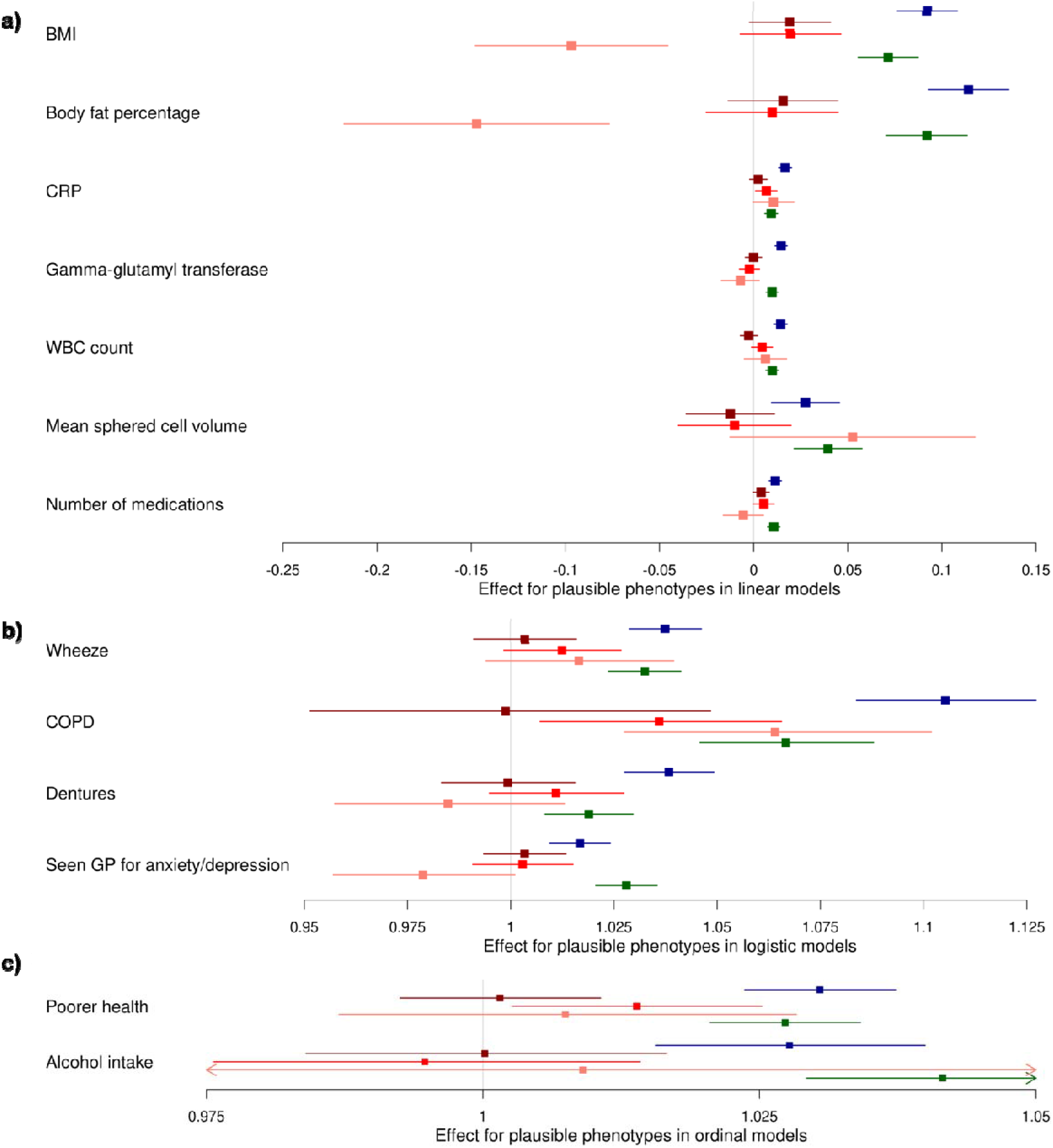
Associations between the PRS for lifetime smoking score (blue), smoking heaviness (red, from darkest to lightest is never, former and current) and smoking initiation (green) and phenotypes plausibly caused by smoking in UK Biobank

Associations between polygenic risk scores for lifetime smoking index (blue), smoking heaviness (red, from darkest to lightest is never, former and current) and smoking initiation (green) and phenotypes plausibly caused by smoking. The figure is split by the type of model used in the analysis; a) linear regression, b) logistic regression, c) ordinal regression. The effect estimate is beta for linear regressions and odds ratios for logistic and ordinal regressions. BMI=body mass index, CRP=C-reactive protein, WBC=white blood cell, COPD=chronic obstructive pulmonary disease, GP=general practitioner.

#### Phenotypes not plausibly caused by smoking

Results from analyses between each of the three smoking-related PRS and the phenotypes not plausibly caused by smoking are shown in Fig 4 and Supplementary Table S8. We found evidence of associations between the PRS for lifetime smoking and smoking initiation and most of our phenotypes not plausibly caused by smoking (p-values=1.06×10^-29^ to 2.77×10^-03^ for lifetime smoking and p-values=1.82×10^-18^ to 4.48×10^-04^ for smoking initiation), with the direction of effect consistent for both PRS. However, we did not observe evidence of any association of back pain experienced in the last month with either of the PRS, or of having ever had an operation on the left-side of the body with the lifetime smoking PRS. The strongest associations were generally observed with the lifetime smoking PRS and the direction of effect was generally the same, where we found associations, with each of the smoking related PRS. For the smoking heaviness PRS, we found positive associations for never smokers with the Townsend deprivation index (b=0.02; 95% CI: 0.005 to 0.03; p=0.007) and liking for cabbage (OR=1.01; 95% CI: 1.00 to 1.03; p=0.02), and for current smokers a negative association with age at first live birth (b=-0.09; 95% CI: −0.19 to −0.003; p=0.04).

**Fig 4.**
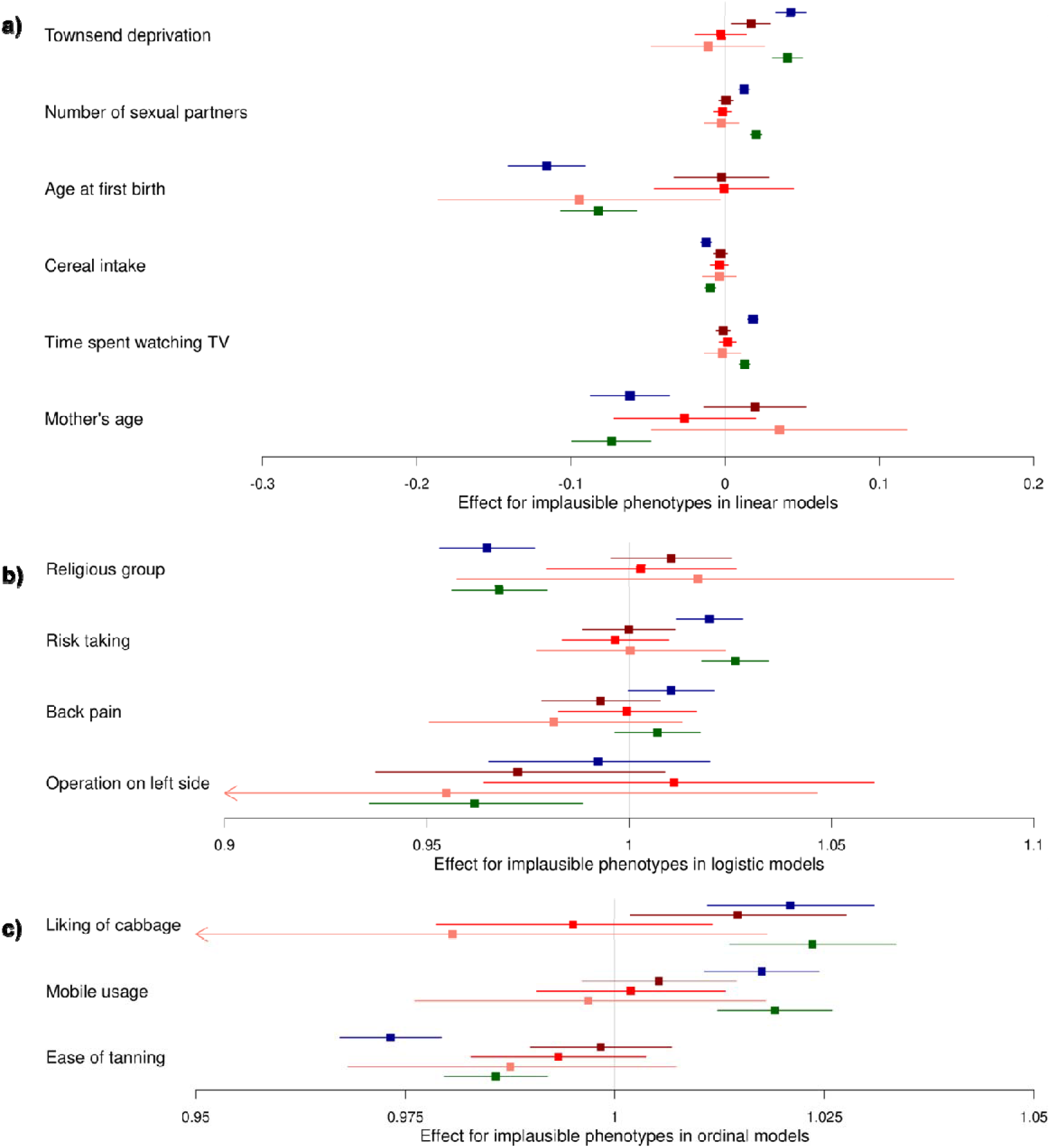
Associations between the polygenic risk scores for lifetime smoking score (blue), smoking heaviness (red, from darkest to lightest is never, former and current) and smoking initiation (green) and phenotypes not plausibly caused by smoking in UK Biobank

Associations between polygenic risk scores for lifetime smoking index (blue), smoking heaviness (red, from darkest to lightest is never, former and current) and smoking initiation (green) and phenotypes not plausibly caused by smoking. The figure is split by the type of model used in the analysis; a) linear regression, b) logistic regression, c) ordinal regression. The effect estimate is beta for linear regressions and odds ratios for logistic and ordinal regressions.

### ALSPAC sample characteristics

Sample characteristics for ALSPAC are shown in Supplementary Table S9. Mean age at the first time point was 28 years (SD=5) for mothers, 31 years (SD=6) for fathers/partners and 5 weeks (SD=3.14) for children. In children 43% were female. Overall, 51% mothers and 55% of fathers/partners had ever smoked. Of those that smoked, the cigarettes per day category with the highest percentage was 10 to 14 for mothers and 20 to 24 for fathers/partners.

### Polygenic risk score associations in ALSPAC (analysis phase)

#### Phenotypes plausibly caused by smoking

Results from analyses between each of the three smoking-related PRS and the phenotypes plausibly caused by smoking are shown in Supplementary Figs S1-S3 and Supplementary Table S10. Compared to UK Biobank, where we found associations of PRS with most of our phenotypes plausibly caused by smoking, we found fewer associations of PRS with those phenotypes in ALSPAC. We did find some evidence of associations between PRS and both increased BMI and body fat percentage, similar to our UK Biobank findings. Specifically, for lifetime smoking we found associations with mother’s (b=0.22; 95% CI: 0.04 to 0.39; p=0.02) and father’s BMI (b=0.23; 95% CI: 0.007 to 0.46; p=0.04), with weaker evidence for child BMI. We found evidence of an association with child’s body fat percentage (b=0.25; 95% CI: 0.02 to 0.48; p=0.03), this was not evident for mothers or fathers/partners, but the direction and size of effect were similar. Furthermore, for lifetime smoking we found positive associations with wheeze (OR=1.07; 95% CI: 1.01 to 1.15; p=0.03) and having seen GP for depression in mothers (T2) (OR=1.09; 95% CI: 1.02 to 1.16; p=0.01), which we also observed in UK Biobank. We found negative associations with mother’s overall health (T1) (OR=0.95; 95% CI: 0.90 to 1.00; p=0.05), father’s overall health (T2) (OR=0.89; 95% CI: 0.80 to 1.00; p=0.05) and child anxiety (OR=0.94; 95% CI: 0.89 to 1.00; p=0.04). This latter finding is the opposite direction to what we observe in UK Biobank.

Unlike in UK Biobank, where we did not find evidence of associations with smoking heaviness PRS in never smokers, in ALSPAC we found a negative association between smoking heaviness PRS in never smokers and number of medications taken in mothers (T2) (b=-0.05; 95% CI: −0.09 to −0.008; p=0.02). With ever smokers, we found a positive association with CRP (b=0.07; 95% CI: 0.01 to 0.14; p=0.02), similar to in UK Biobank, and negative associations with alcohol consumption in mothers (OR=0.92; 95% CI: 0.86 to 0.98; p=0.006) and number of medications in fathers/partners (T1) (b=-0.14; 95% CI: −0.24 to −0.03; p=0.01). However, we did not replicate our UK Biobank associations with COPD, overall health, BMI and body fat percentage in ALSPAC.

Finally, for the smoking initiation PRS we found positive associations with BMI (b=0.36; 95% CI: 0.19 to 0.54; p=0.00005) and body fat percentage in mothers (b=0.34; 95% CI: 0.05 to 0.62; p=0.02). Again, the direction of effect was the same across all samples for these and the same direction as that in UK Biobank. We also found a negative association with child anxiety (OR=0.92; 95% CI: 0.87 to 0.97; p=0.004), in the opposite direction to UK Biobank, and a positive association with alcohol consumption in mothers (OR=1.06; 95% CI: 1.02 to 1.11; p=0.006), similar to in UK Biobank. However, we did not replicate other findings with phenotypes plausibly caused by smoking that we found in UK Biobank.

#### Phenotypes not plausibly caused by smoking

Results from analyses between each of the three smoking-related PRS and the phenotypes not plausibly caused by smoking are shown in Supplementary Figs S4-S6 and Supplementary Table S11. Similarly to in UK Biobank, we found many associations between PRS for lifetime smoking and phenotypes not plausibly caused by smoking. For lifetime smoking PRS we found positive associations with Townsend deprivation index in mothers (both timepoints) (T1: OR=1.06; 95% CI: 1.01 to 1.12; p=0.02 and T2: OR=1.05; 95% CI: 1.00 to 1.09; p=0.05), time spent watching TV in mothers on weekdays (b=0.05; 95% CI: 0.02 to 0.08; p=0.0006) and weekend days (b=0.03; 95% CI: 0.006 to 0.06; p=0.02) and cabbage intake in children (OR=1.06; 95% CI: 1.01 to 1.11; p=0.01), and negative associations with mother’s age at first pregnancy (b=-0.19; 95% CI: −0.28 to −0.10; p=5.85×10^-05^), attending a place of worship in mothers (OR=0.91; 95% CI: 0.87 to 0.96; p=0.0002) and children (OR=0.90; 95% CI: 0.85 to 0.95; p=0.00008), reduced wholegrain cereal consumption (both timepoints) (T1: OR=0.92; 95% CI: 0.86 to 0.96; p=0.0003 and T2: OR=0.94; 95% CI: 0.89 to 0.98; p=0.007) and increased intake of other cereals in mothers (T1) (OR=1.05; 95% CI: 1.00 to 1.09; p=0.04), avoiding risks in children (OR=0.92; 95% CI: 0.88 to 0.96; p=0.0005) and age of the father’s mother at time of questionnaire i.e., lower age (b=-0.17; 95% CI: −0.32 to −0.03; p=0.02).

We did not find any associations with smoking heaviness PRS in never smokers in all of our samples compared to the few we found in UK Biobank – potentially suggesting less pleiotropy for these variants. With ever smokers we found an association between smoking heaviness PRS and attending a place of worship less frequently in mothers (OR=0.89; 95% CI: 0.83 to 0.96; p=0.003), which we did not find in UK Biobank.

Finally, similarly to the many associations we found in UK Biobank between the smoking initiation PRS and phenotypes not plausibly caused by smoking, in ALSPAC for the smoking initiation PRS we found positive associations with cabbage intake in children (OR=1.05; 95% CI: 1.01 to 1.11; p=0.02) and mobile phone usage in mothers (OR=1.08; 95% CI: 1.00 to 1.16; p=0.05), and negative associations with age at first pregnancy in mothers (b=-0.15; 95% CI: −0.24 to −0.06; p=0.001), attending a place of worship in mothers (OR=0.92; 95% CI: 0.88 to 0.96; p=0.0004) and children (OR=0.95; 95% CI: 0.90 to 1.00; p=0.05), oat cereal intake in mothers (T2) (OR=0.94; 95% CI: 0.89 to 0.99; p=0.01), avoiding risks in children (both timepoints) (T1: OR=0.93; 95% CI: 0.88 to 0.97; p=0.001 and T2: OR=0.95; 95% CI: 0.91 to 1.00; p=0.04) and mother’s (b=-0.16; 95% CI: −0.30 to −0.02; p=0.03) and age of the father’s mother at time of questionnaire i.e., lower age (b=-0.35; 95% CI: −0.67 to −0.03; p=0.03).

## Discussion

We found evidence of likely horizontal pleiotropy for genetic risk scores intended to capture cigarette smoking phenotypes. This has implications for any study using these exposures within an MR framework, and suggests caution may also be required for studies of other complex behaviour phenotypes.

Specifically, we found evidence that PRS for lifetime smoking index and smoking initiation were associated with most phenotypes not plausibly caused by smoking (as well as all phenotypes plausibly caused by smoking) in UK Biobank. There was less evidence of associations with the smoking heaviness PRS, but the strongest associations were found for current smokers. Some (although not all) of these results were replicated in ALSPAC. Our results are in line with recent studies suggesting associations of increased smoking initiation PRS with increased risk-taking behaviours and decreased age at first birth, amongst other phenotypes [12, 13, 22]. However, we also found novel associations with other phenotypes not plausibly caused by smoking, suggesting that these potential pleiotropic effects may occur via a range of pathways.

It is difficult to know how associations with phenotypes not plausibly caused by smoking might arise; they could be a result of correlated pleiotropy, uncorrelated pleiotropy, or mis-specification of the primary phenotype. For example, the presence of a mis-specified, unknown, primary phenotype on a vertical pleiotropic pathway may induce correlated pleiotropy. It is unlikely that an unknown phenotype is actually on the causal pathway from exposure to outcome because we consider the outcomes examined to be not plausibly caused by smoking. The presence of pleiotropy is further supported by evidence of associations in never smokers and children, where these phenotypes cannot be downstream of smoking because there has been no first-hand exposure to smoking.

The fact we find less evidence of associations for the smoking heaviness PRS fits with the theory that smoking initiation measures in particular may be capturing other underlying phenotypes. The smoking heaviness PRS is more likely to capture biological function such as nicotinic pathways, as demonstrated in a previous PheWAS study in UK Biobank, where associations were observed with poorer lung function, higher blood assay levels, COPD, emphysema, cancer, and greater facial aging [26] among ever smokers. Whereas the smoking initiation measures are more likely to capture other phenotypes as well, as demonstrated in this study with our phenotypes not plausibly caused by smoking.

UK Biobank has some limitations in terms of selection bias. Compared to respondents of the same age range in national surveys, participants in UK Biobank were more likely to be homeowners, with lower BMIs, less likely to be current smokers and to drink alcohol daily, suffer from fewer self-reported health conditions and have lower mortality in follow-up [27]. This indicates a ‘healthy volunteer’ effect in UK Biobank which should be considered when interpreting our results. Therefore, we also conducted analyses in ALSPAC, which may have different patterns of selection due to one being a cross-sectional study and the other being a birth cohort. We found that some effects were consistent across cohorts. However, for other relationships, effects were not consistent between the cohorts. For example, for smoking heaviness PRS in ever smokers we found that a greater PRS was associated with less frequent attendance at a place of worship in ALSPAC, but we did not find any association in UK Biobank. We also found associations with other outcomes in UK Biobank but not in ALSPAC, although the direction of effect was often consistent. These differences could be due to different patterns in selection, different phenotype definitions, different sample sizes, or they could be due to Winner’s Curse as UK Biobank was our discovery sample and ALSAPC was our replication sample [28].

It is also worth noting that we found an association between increased smoking heaviness PRS and fewer medications being taken in mothers who had never smoked. This is unexpected given that number of medications is one of our phenotypes plausibly caused by smoking, although this could reflect low health seeking behaviour. This further highlights that horizontal pleiotropic pathways can exist even for phenotypes plausibly downstream of smoking.

Finally, it is worth discussing our findings in children for our negative control analyses where we also replicated findings; for example, we found a negative association with avoiding taking risks, suggesting that lifetime smoking and smoking initiation PRS may pick up risk-taking behaviours even in children who are unlikely to have considered smoking. This supports previous findings [12, 13] and further supports our conclusions that there is horizontal pleiotropy in smoking PRS which incorporate smoking initiation. Data from children could be included in additional MR analyses, as these would be conducted in individuals after the age where smoking is likely to have been initiated, to help examine these associations further.

A few limitations should be noted when interpreting our results. Firstly, a sub-sample of the UK Biobank cohort were part of the UK Biobank Lung Exome Variant Evaluation (BiLEVE) study [29], which oversampled for smokers and for which a different genotyping chip was used compared to the rest of the sample, which could introduce collider bias into our results. It may also be the case that smoking phenotypes are under reported, for example, those identifying as ‘social smokers’ may underestimate their smoking heaviness or may not respond as being a smoker. However, we used three different measures of smoking so this limitation may be countered partially by that, and this is likely to only be the case for a small number of individuals. In addition, our analyses were conducted in two European ancestry samples, therefore, our results may be less generalisable to other ancestry groups. Also, some of the phenotypes plausibly caused by smoking we identified have potentially complex relationships with smoking, for example, alcohol consumption and mental health. This may mean that there are bidirectional relationships, for example. However, given the evidence of associations between smoking and these phenotypes we would argue that they are still valid phenotypes plausibly caused by smoking. It may also be the case that there are alternative explanations for our findings. For example, population stratification could account for some of the associations we find, although we adjusted for PCs to account for this. Furthermore, our findings in children prior to smoking could also potentially be due to dynastic effects, whereby the parents share a genetic propensity to smoke with their child, and thus second-hand smoke exposure from their parents could induce an association between the child’s genotype and a given outcome. Finally, collider bias could also impact our results, although previous simulations suggest that in such a large sample any collider bias from the effect of the smoking heaviness PRS on smoking status is unlikely to have a large impact on the effect estimate [26].

We found evidence of horizontal pleiotropy in exposures frequently included in MR analyses. Specifically, our results suggest that the SNPs used as genetic instruments for smoking related phenotypes may in fact be instrumenting other phenotypes. This has important implications for any study using smoking-related SNPs or SNPs for other complex traits as instruments in an MR approach, as our inferences from these studies may be incorrect if this is not considered. Our results are particularly important to consider especially given that as GWAS become increasingly larger; for complex traits in particular, it is likely that the problem of pleiotropy will also become more widespread, as discussed elsewhere [5]. Therefore, methods designed to address issues around pleiotropy (e.g., MR-Egger, MR-PRESSO, and GSMR for MR analyses) should be included in genetically-informed causal inference analyses. Researchers should also consider other approaches to address this potential pleiotropy, for example, including outcomes not plausibly caused by an exposure of interest and negative control analyses e.g., including unexposed groups. We found evidence of horizontal pleiotropy to a lesser extent for smoking heaviness, suggesting that for analyses with smoking exposures, SNPs for smoking heaviness may be better suited, especially when including a negative control analysis of non-smokers or children prior to smoking onset to assess possible bias from horizontal pleiotropy. However, caution is still advised when using these instruments and consideration of how this may impact results should be given on a study-by-study basis. We recommend that similar analyses are also conducted for studies with other complex traits as well.

## Statements and Declarations

### Author contributions

MRM, GDS and REW contributed to the study conception and design. Data preparation and analysis were performed by ZER and TGR. The first draft of the manuscript was written by ZER, REW and JNK. All authors commented on subsequent versions of the manuscript. All authors read and approved the final manuscript.

### Ethics approval

UK Biobank received ethics approval from the Research Ethics Committee (REC reference for UK Biobank is 11/NW/0382). Ethics approval was obtained from the ALSPAC Ethics and Law Committee and the Local Research Ethics Committees (http://www.bristol.ac.uk/alspac/researchers/research-ethics/).

### Consent to participate

For UK Biobank informed consent was obtained from all individual participants at the point of data collection. For ALSPAC consent for biological samples was collected in accordance with the Human Tissue Act (2004). Informed consent for the use of data collected via questionnaires and clinics was obtained from participants following the recommendations of the ALSPAC Ethics and Law Committee at the time.

## Data Availability

GWAS data for smoking initiation with UK Biobank and 23andMe removed can be found here: https://conservancy.umn.edu/handle/11299/201564. Full GWAS summary statistics for the 23andMe discovery data set (which we combined with the publicly available smoking initiation data) will be made available through 23andMe to qualified researchers under an agreement with 23andMe that protects the privacy of the 23andMe participants. Please visit https://research.23andme.com/collaborate/#dataset-access/ for more information and to apply to access the data.
ALSPAC data access is through a system of managed open access (http://www.bristol.ac.uk/alspac/researchers/access/).
UK Biobank data are available through a procedure described at http://www.ukbiobank.ac.uk/using-the-resource/.

## Acknowledgements

This research has been conducted using data from UK Biobank (project ID: 16729), a major biomedical database (http://www.ukbiobank.ac.uk/). We are extremely grateful to all the families who took part in the ALSPAC study, the midwives for their help in recruiting them, and the whole ALSPAC team, which includes interviewers, computer and laboratory technicians, clerical workers, research scientists, volunteers, managers, receptionists and nurses. We thank all the contributors to the consortia we have used GWAS results from in our analyses. We would like to thank the research participants and employees of 23andMe, inc. for making this work possible.

