## Supplementary Materials for "Exploring pleiotropy in Mendelian randomisation analyses: What are genetic variants associated with “cigarette smoking initiation” really capturing?"

European Journal of Epidemiology

Zoe E. Reed[1,2*], Robyn E. Wootton[1,2,3], Jasmine N. Khouja[1,2], Tom G. Richardson[2], Eleanor Sanderson[2], George Davey Smith[2], Marcus R Munafò[1,2,4]

1 School of Psychological Science, University of Bristol, Bristol, UK

2 MRC Integrative Epidemiology Unit at the University of Bristol, Bristol, UK

3 Nic Waals Institute, Lovisenberg Diaconal Hospital, Oslo, Norway

4 National Institute for Health Research Bristol Biomedical Research Centre, University Hospitals Bristol NHS Foundation Trust and University of Bristol, Bristol, UK

**Contents**

| | |
|---|---|
| *Section 1. Smoking related exposures* | *Page 4* |
| *Section 2. PheWAS to identify plausible and implausible phenotypes* | *Page 4* |
| *Section 3. Plausible phenotype selection* | *Page 5* |
| *Section 4. UK Biobank genetic data* | *Page 6* |
| *Section 5. ALSPAC genetic data* | *Page 6* |
| *Section 6. Genome-wide association studies in UK Biobank* | *Page 8* |
| *Section 7. Polygenic risk score construction in UK Biobank* | *Page 8* |
| *Section 8. Polygenic risk score construction in ALSPAC* | *Page 8* |
| *Supplementary Table S1. Top 100 associaitons in the PheWAS* | *Page 9* |
| *Supplementary Table S2. Details of plausible phenotypes in UK Biobank.* | *Page 13* |
| *Supplementary Table S3. Details of implausible phenotypes in UK Biobank* | *Page 15* |
| *Supplementary Table S4. ALSPAC phenotypes for plausible phenotypes* | *Page 17* |
| *Supplementary Table S5. ALSPAC phenotypes for implausible phenotypes* | *Page 22* |
| *Supplementary Table S6. Summary statistics for UK Biobank sample with data available* | *Page 27* |
| *Supplementary Table S7. Associations between the polygenic risk scores for lifetime smoking score, smoking heaviness and smoking initiation and plausible outcomes in UK Biobank* | *Page 29* |
| *Supplementary Table S8. Associations between the polygenic risk scores for lifetime smoking score, smoking heaviness and smoking initiation and implausible outcomes in UK Biobank* | *Page 32* |
| *Supplementary Table S9. Summary statistics for ALSPAC sample with data available* | *Page 35* |
| *Supplementary Table S10. Associations between the polygenic risk scores for lifetime smoking score, smoking heaviness and smoking initiation and plausible outcomes in ALSPAC* | *Page 42* |
| *Supplementary Table S11. Associations between the polygenic risk scores for lifetime smoking score, smoking heaviness and smoking initiation and implausible outcomes in ALSPAC* | *Page 49* |

*Supplementary Fig S1. Associations between polygenic risk scores for lifetime smoking score (blue), smoking heaviness (red, from darkest to lightest is never, former and current) and smoking initiation (green) and plausible phenotypes (linear models)* *Page 55*

*Supplementary Fig S2. Associations between polygenic risk scores for lifetime smoking score (blue), smoking heaviness (red, from darkest to lightest is never, former and current) and smoking initiation (green) and plausible phenotypes (logistic models)* *Page 57*

*Supplementary Fig S3. Associations between polygenic risk scores for lifetime smoking score (blue), smoking heaviness (red, from darkest to lightest is never, former and current) and smoking initiation (green) and plausible phenotypes (ordinal models)* *Page 58*

*Supplementary Fig S4. Associations between polygenic risk scores for lifetime smoking score (blue), smoking heaviness (red, from darkest to lightest is never, former and current) and smoking initiation (green) and implausible phenotypes (linear models)* *Page 60*

*Supplementary Fig S5. Associations between polygenic risk scores for lifetime smoking score (blue), smoking heaviness (red, from darkest to lightest is never, former and current) and smoking initiation (green) and implausible phenotypes (logistic models)* *Page 61*

*Supplementary Fig S6. Associations between polygenic risk scores for lifetime smoking score (blue), smoking heaviness (red, from darkest to lightest is never, former and current) and smoking initiation (green) and implausible phenotypes (ordinal models)* *Page 62*

*References* *Page 63*

### *Section 1. Smoking related exposures*

In UK Biobank, for smoking initiation, we included any participants who indicated they currently or previously smoked in the ever smoked group and anyone who indicated they had never smoked in the other group (UKBB field number: 20116). For smoking heaviness, we used number of cigarettes smoked per day (UKBB field number: 3456). Where the participant was a former smoker, cigarettes per day when they did smoke was used as a measure of heaviness (UKBB field number: 2887). Participants who reported smoking less than one or more than 150 cigarettes per day were removed by UK Biobank and those reporting more than 100 cigarettes per day were asked to confirm this. Participants were informed that for hand-rolled cigarettes, one gram of tobacco was equivalent to one cigarette.

In UK Biobank, the lifetime smoking index, which was constructed in a previous study, was designed to capture a number of aspects of smoking behaviour (i.e., initiation, heaviness and duration). We used measures of smoking status (current, former or never), age at initiation (UKBB field number: 3436 and 2867) and cessation (UKBB field number: 2897), if applicable, and number of cigarettes smoked per day (as for our smoking heaviness measure). The smoking measures were combined with a simulated half-life constant which captured the exponentially decreasing effect of smoking over time on health outcomes. The best fitting value for this was 18 and further details of the simulations to obtain this value can be found in the original article describing this measure [1]. We standardised the lifetime smoking index to have a mean of 0 and a standard deviation (SD) of 1.

Data on smoking status and cigarettes per day (CPD) were also available in ALSPAC. Smoking status was collected for both mothers and fathers/partners when they were asked if they had ever been a smoker (collected during pregnancy). We included measures of cigarettes per day obtained at two timepoints for mothers and one for fathers/partners. For mothers we included two timepoints as the first measure was obtained just after pregnancy and may not be a good representation of usual smoking habits for this reason. Therefore, we also included a measure collected 8 months after birth. For the first measure mothers were asked how many cigarettes they smoked per day over the past week (categorical variable with 'not at all', '1 to 4', '5 to 9', '10 to 14', '15 to 19', '20 to 24', '25 to 29' and '30 or more'). For the second measure mothers were asked how many cigarettes per day they currently smoke (same categories as previous). The fathers/partners CPD measure was also obtained 8 months after birth, and they were asked the same question as mothers.

### *Section 2. PheWAS to identify plausible and implausible phenotypes*

We initially conducted a phenome-wide association study (PheWAS) [2] for smoking initiation using a polygenic risk score of smoking initiation as the exposure, constructed in UK Biobank. To avoid sample overlap, we used GWAS summary statistics from the GWAS and Sequencing Consortium of Alcohol and Nicotine use (GSCAN) GWAS [3] for smoking initiation, excluding the UK Biobank sample. We meta-analysed results from GWAS of 23andMe, Inc. only data and all results excluding UK Biobank and 23andMe. The meta-analysis was conducted using the genome-wide association meta-analysis (GWAMA) software [4]. We used genome-wide significant SNPs only in our polygenic risk score. The PheWAS was conducted using the PHEnome Scan ANalysis Tool (PHESANT) software package [5], which performs phenome scans on data from UK Biobank. We used this to test the association of our polygenic risk score with all of these outcomes (21,409 variables). Of these, 566 variables were associated with the polygenic risk score (at a Bonferroni adjusted p-value threshold of $2.34 \times 10^{-06}$). From the top 100 most associated of these (a threshold we decided *a priori*) we selected variables to be plausible and implausible phenotypes in these analyses (our outcome variables). We did not include those that were conceptually related to the main smoking phenotypes e.g., age stopped smoking and for similar variables we selected the one that we believed captured the most information. Plausible phenotypes were those known (or strongly believed) to be causally related to smoking. Implausible phenotypes are those that are unlikely to be causally related to smoking.

### *Section 3. Plausible phenotype selection*

Plausible phenotypes are those likely to be caused by smoking, whereas implausible phenotypes are those unlikely to be caused by smoking. We have listed these below with a brief description of the evidence for why a phenotype is considered plausibly downstream of smoking.

- Body mass index (BMI): There is evidence that smoking leads to a decrease in BMI [6–8], further supported by studies that demonstrate an increase in BMI following smoking cessation [9].
- Body fat percentage: There is less evidence for the relationship with body fat percentage, however associations tend to be in the same direction as BMI (i.e., smoking is associated with lower body fat percentage) [10]. As there is overlap between BMI and body fat percentage, we believe that body fat percentage is also plausibly downstream of smoking.
- Wheeze: Previous studies have found an association between smoking and experiencing wheezing [11]. Wheeze is also a core symptom of asthma and smoking may causally lead to an increased risk of asthma [12].
- C-reactive protein (CRP): CRP levels have been found to be increased in smokers [13], also observed in a Mendelian Randomisation study [14].
- Ever reported COPD: Smoking has been well-established as a leading cause of COPD [15].
- Had dentures: Smoking is associated with poorer oral health, including tooth loss [16,17], which would in turn lead to increased use of dentures.
- Overall health rating: In line with some of the other evidence of the negative health impact of smoking presented in this section, smoking impacts a range of health-related outcomes and is associated with poorer overall health [18].
- Gamma glutamyl transferase (GGT): GGT has been found to be increased in current and former smokers [19].
- White blood cell count: There is evidence that smoking leads to increased counts of white blood cells [20,21].
- Mean sphered cell volume: Smoking is associated with increased red blood cell volume [22].
- Seen GP for nerves, anxiety, or depression: There is evidence to suggest that smoking may increase risk of both anxiety and depression [1,23–25], although a bidirectional effect may also be present.

- Numbers of medications taken: We did not find studies specifically examining this relationship. However, given that smoking is associated with poorer health, increased medication use will likely be downstream of this effect as well.
- Alcohol consumption: There is evidence to suggest that smoking could lead to increased alcohol consumption, but this relationship is likely to be complex [26].

***Section 4. UK Biobank genetic data***

There were 488,377 participants with genotyped samples of which 49,979 were genotyped using the UK BiLEVE array and 438,398 using the UK Biobank axiom array. Pre-imputation quality control, phasing and imputation have been described elsewhere [27]. In summary, multiallelic SNPs and those with a MAF≤1% were removed. Phasing of genotype data was performed using a modified version of the SHAPEIT2 algorithm. The SNPs used were imputed to the Haplotype Reference Consortium (HRC) reference panel, using IMPUTE2 algorithms. A graded filtering with different imputation qualities for different MAF ranges was used (Info>0.3 for MAF>3%, Info>0.6 for MAF 1-3%, Info>0.8 for MAF 0.5-1% and Info>0.9 for MAF 0.1-0.5%), where MAF and info scores were recalculated on an in-house derived 'European' subset. Individuals with sex-mismatch or sex-chromosome aneuploidy were excluded (N=814). In-house quality control filtering of the UK Biobank data is described in a published protocol [28].

We restricted the sample to individuals of "White British" ancestry and who have very similar ancestral backgrounds according to principal components analysis (N=409,703) [27]. Estimated kinship coefficients using the KING toolset [29] identified 107,162 pairs of related individuals. An in-house algorithm was then applied to this list and preferentially removed the individuals related to the greatest number of other individuals until no related pairs remain. These individuals were excluded (N=79,448). Additionally, 2 individuals were removed due to them relating to a very large number (>200) of individuals. After these exclusions and excluding individuals who had withdrawn their data, we included 336,988 individuals in our analyses.

***Section 5. ALSPAC genetic data***

Samples for children were genotyped using the Illumina HumanHap 550 quad chip. Genome-wide data for children were generated by Sample Logistics and Genotyping Facilities at the Wellcome Trust Sanger Institute and LabCorp (Laboratory Corporation of America) using support from 23andMe. Quality control measures were used, and individuals were excluded based on gender mismatches, minimal or excessive heterozygosity, disproportionate levels of individual missingness (>3%) and insufficient sample replication (identity by descent [IBD] <0.8). Population stratification was assessed by multidimensional scaling analysis and compared with Hapmap II (release 22) European descent (CEU), Han Chinese, Japanese and Yoruba reference populations; all individuals with non-European ancestry were removed. SNPs with a minor allele frequency (MAF) of <1%, a call rate of <95% or evidence of violations of Hardy-Weinberg equilibrium ($P<5x10^{-07}$) were removed. Cryptic relatedness was measured as the proportion of IBD>0.1.

Samples for mothers were genotyped using the Illumina human660W-quad array at Centre National de Génotypage (CNG) and genotypes were called with Illumina GenomeStudio.

Quality control measures were used. SNPs were removed if they displayed more than 5% missingness and a Hardy-Weinberg equilibrium P value of $<1x10^{-06}$. SNPs with a MAF <1% were removed. Samples were excluded if they displayed >5% missingness, had indeterminable X chromosome heterozygosity or extreme autosomal heterozygosity. Samples showing evidence of population stratification were identified by multidimensional scaling of genome-wide identity by state pairwise distances using the four HapMap populations as reference, and then excluded. Cryptic relatedness was assessed using an IBD estimate >0.125, which is expected to correspond to approximately 12.5% alleles shared IBD or a relatedness at the first cousin level.

Related individuals that passed all other quality control thresholds were retained during subsequent phasing and imputation. 9,115 children and 500,527 SNPs and 9.048 mothers and 526,688 SNPs passed these filters. 477,482 SNP genotypes in common between mothers and children were removed. SNPs with genotype missingness >1% were also removed. 321 individuals were removed due to potential ID mismatches. Haplotypes were estimated using ShapeIT (V2.r644) which utilises relatedness during phasing. A phased version of the 1000 genomes reference panel (Phase 1, version 3) was obtained from the Impute2 reference data repository (phased using ShapeIT v2.r644, haplotype release date Dec 2013). Imputation of the target data was performed using Impute V2.2.2 against the reference panel using all 2186 reference haplotypes (including non-Europeans).

Samples for fathers/partners were genotyped using the Illumina HumanCoreExome chip genotyping platforms by the ALSPAC lab and called using GenomeStudio. Quality control measures were used. SNPs were removed if they displayed a call rate of <95%, evidence for violations of Hardy-Weinberg equilibrium ($P<1x10^{-07}$), failed GenomeStudio quality control measures or were duplicates. Samples were excluded if they displayed >5% missingness, had minimal or excessive heterozygosity, gender mismatches or possible contamination. Samples showing evidence of population stratification were identified by multidimensional scaling analysis and compared with 1000 Genomes phase 3 data and principal component analysis. All individuals with non-European ancestry were removed. Cryptic relatedness was assessed in GCTA using relatedness >0.1.

Data that passed quality control steps were phased. 155,336 monomorphic SNPs, 1033 markers not in 1000 genomes, 11,842 A/T or G/C SNPs and 10 duplicate sites were then removed to give 337,732 SNPs on chromosomes 1-23. Of the 329,363 markers on chromosomes 1-22, 298,742 overlapped the reference genome. Data was imputed to the 1000 genomes phase 1 version 3 using the Michigan Imputation Server. Individuals were also removed if their sample ID assigned historically did not match the genetically assigned sample ID.

There were 8,237 eligible children, 8,196 eligible mothers and 2,201 fathers/partners with available genotype data after exclusion of related subjects and these quality control steps.

### *Section 6. Genome-wide association studies in UK Biobank*

We conducted genome-wide association studies (GWAS) of smoking heaviness (cigarettes per day) and smoking initiation (ever/never) using the BOLT-LMM software [30], adjusting for age, sex and genotyping chip in the model. BOLT-LMM performs a linear regression and therefore for our binary outcome of smoking initiation the output betas represented an absolute 'risk difference' scale. Therefore, we transformed the betas and standard errors to obtain log odds ratios using following formula (for standard error we replace beta with standard error):

$$logOR = \frac{\beta bolt}{\mu(1-\mu)}$$

where $\mu$ is the case prevalence ($ncase/(ncase + ncontrol)$). We used the number of cases and controls in our sample to obtain a prevalence of 0.45.

***Section 7. Polygenic risk score construction in UK Biobank***

To construct the polygenic risk scores we identified genetic variants robustly associated with each smoking trait (based on $P<5x10^{-8}$) and then pruned them to identify independent genetic variants by applying linkage disequilibrium (LD) clumping (based on $r^2<0.001$). A reference panel of 10,000 randomly selected and unrelated UK Biobank individuals of European descent was used for LD clumping [31]. Polygenic risk scores were subsequently generated using the software PLINK [32] and standardised to have a mean of 0 and an SD of 1.

***Section 8. Polygenic risk score construction in ALSPAC***

We created polygenic risk scores for smoking heaviness, smoking initiation and lifetime smoking in ALSPAC. For smoking heaviness and initiation, we used summary statistics from the GWAS we conducted in UK Biobank. For lifetime smoking we used summary statistics from a published GWAS of lifetime smoking [1]. We filtered SNPs by an imputation score of 0.8. After this filtering and selecting SNPs where genotype data was available in our ALSPAC samples, SNPs were clumped for linkage disequilibrium, using the –clump command in PLINK and an $R^2$ of 0.001. We generated weighted polygenic risk scores for each phenotype using the 'score' command in PLINK which creates averages of valid per-allele scores, using the weights from the respective GWAS (effect estimates for smoking heaviness and lifetime smoking and log odds ratios for smoking initiation). We present results for polygenic risk scores constructed at the $p <5x10^{-8}$ threshold in the discovery GWAS. Polygenic risk scores were z-standardised; therefore, results can be interpreted as per standard deviation (SD) increase in score.

For the lifetime smoking polygenic risk score there were approximately 142 SNPs, for the smoking heaviness polygenic risk score there were approximately 9 SNPs and for the smoking initiation polygenic risk score there were approximately 151 SNPs.

**Supplementary Table S1. Top 100 associaitons in the PheWAS**

| Phenotype | N (total or no/yes) | Beta | Lower CI | Upper CI | P-value |
|---|---|---|---|---|---|
| Past tobacco smoking | 311147 | -0.14 | -0.15 | -0.13 | $<2.06x10^{-269}$ |
| Smoking status | 184005/335804 | -999.00 | -999.00 | -999.00 | $<2.06E^{-269}$ |
| Ever smoked | 132831/203010 (335841) | 0.14 | 0.13 | 0.14 | $<2.06E^{-269}$ |
| Age first had sexual intercourse | 296920 | -0.06 | -0.07 | -0.06 | $2.06E^{-269}$ |
| Current tobacco smoking | 336812 | 0.17 | 0.16 | 0.18 | $1.67E^{-193}$ |
| Maternal smoking around birth | 201188/88460 (289648) | 0.11 | 0.10 | 0.11 | $3.41E^{-148}$ |
| Tobacco smoking | 51588/85937 | -999.00 | -999.00 | -999.00 | $7.28E^{-143}$ |
| Date F17 first reported (mental and behavioural disorders due to use of tobacco) | 311833/25155 (336988) | 0.15 | 0.14 | 0.16 | $1.11E^{-113}$ |
| Qualifications: College or University degree | 227110/106749 (333859) | -0.08 | -0.09 | -0.08 | $8.45E^{-109}$ |
| Lifetime number of sexual partners | 277634 | 0.08 | 0.07 | 0.08 | $1.17E^{-98}$ |
| Leg fat percentage (right) | 331063 | 0.02 | 0.02 | 0.02 | $1.36E^{-93}$ |
| Leg fat percentage (left) | 331045 | 0.02 | 0.02 | 0.02 | $1.87E^{-93}$ |
| Education score (England) | 288592 | 0.04 | 0.03 | 0.04 | $2.33E^{-90}$ |
| Leg fat mass (left) | 331042 | 0.03 | 0.02 | 0.03 | $3.60E^{-90}$ |
| Leg fat mass (right) | 331060 | 0.03 | 0.02 | 0.03 | $7.94E^{-90}$ |
| Body mass index (BMI) | 331083 | 0.03 | 0.03 | 0.04 | $1.00E^{-79}$ |
| Body mass index (BMI) | 335908 | 0.03 | 0.03 | 0.04 | $8.72E^{-79}$ |
| Qualifications: A levels/AS levels or equivalent | 241673/92186 (333859) | -0.07 | -0.08 | -0.07 | $5.57E^{-78}$ |
| Whole body fat mass | 330540 | 0.03 | 0.03 | 0.03 | $1.22E^{-77}$ |
| Employment score (England) | 288592 | 0.03 | 0.03 | 0.04 | $8.03E^{-76}$ |
| Waist circumference | 336415 | 0.03 | 0.02 | 0.03 | $6.41E^{-74}$ |
| Body fat percentage | 330893 | 0.02 | 0.02 | 0.03 | $1.18E^{-71}$ |
| Own or rent accommodation lived in | 182447/332896 | -999.00 | -999.00 | -999.00 | $2.68E^{-70}$ |
| Arm fat mass (left) | 330933 | 0.03 | 0.03 | 0.03 | $6.72E^{-70}$ |
| Arm fat mass (right) | 330992 | 0.03 | 0.03 | 0.03 | $9.63E^{-70}$ |
| Health score (England) | 288592 | 0.03 | 0.03 | 0.04 | $1.80E^{-69}$ |
| Index of Multiple Deprivation (England) | 288592 | 0.03 | 0.03 | 0.04 | $6.44E^{-69}$ |

| | | | | | |
|---|---|---|---|---|---|
| Trunk fat mass | 330861 | 0.03 | 0.03 | 0.03 | 2.98E$^{-67}$ |
| Qualifications | 276778/57081 (333859) | 0.08 | 0.07 | 0.09 | 6.02E$^{-66}$ |
| Age completed full time education | 226242 | -0.07 | -0.07 | -0.06 | 1.18E$^{-63}$ |
| Weight | 331090 | 0.03 | 0.02 | 0.03 | 1.53E$^{-63}$ |
| Weight | 336024 | 0.03 | 0.02 | 0.03 | 3.08E$^{-63}$ |
| Income score (England) | 288592 | 0.03 | 0.03 | 0.03 | 3.42E$^{-63}$ |
| Wheeze or whistling in the chest in last year | 263092/68032 (331124) | 0.07 | 0.06 | 0.08 | 8.03E$^{-63}$ |
| Arm fat percentage (right) | 331017 | 0.02 | 0.02 | 0.02 | 6.41E$^{-62}$ |
| Age at first live birth | 124051 | -0.05 | -0.05 | -0.04 | 4.05E$^{-61}$ |
| Arm fat percentage (left) | 330966 | 0.02 | 0.02 | 0.02 | 2.27E$^{-59}$ |
| Trunk fat percentage | 330880 | 0.02 | 0.02 | 0.03 | 9.88E$^{-57}$ |
| Alcohol usually taken with meals | 55075/117237 (172312) | -0.08 | -0.09 | -0.07 | 3.93E$^{-53}$ |
| C-reactive protein | 320589 | 0.03 | 0.02 | 0.03 | 5.52E$^{-53}$ |
| Townsend deprivation index at recruitment | 336591 | 0.03 | 0.02 | 0.03 | 2.25E$^{-52}$ |
| Date J44 first reported (other chronic obstructive pulmonary disease) | 326520/10468 (336988) | 0.15 | 0.13 | 0.17 | 2.42E$^{-49}$ |
| Mouth/teeth dental problems: Dentures | 279583/56336 (335919) | 0.07 | 0.06 | 0.08 | 4.53E$^{-47}$ |
| Average weekly beer plus cider intake | 241456 | 0.06 | 0.05 | 0.07 | 1.05E$^{-46}$ |
| Light smokers, at least 100 smokes in lifetime | 50401/41710 (92111) | 0.09 | 0.08 | 0.10 | 1.22E$^{-41}$ |
| Hip circumference | 336371 | 0.02 | 0.02 | 0.03 | 5.41E$^{-41}$ |
| Overall health rating | 335806 | 0.05 | 0.04 | 0.05 | 1.04E$^{-40}$ |
| Arm predicted mass (left) | 330914 | 0.01 | 0.01 | 0.02 | 1.44E$^{-40}$ |
| Attendance/disability/mobility allowance | 19082/315780 (334862) | -0.10 | -0.11 | -0.08 | 3.67E$^{-40}$ |
| Arm fat-free mass (left) | 330926 | 0.01 | 0.01 | 0.02 | 1.14E$^{-39}$ |
| Leisure/social activities: Religious group | 288619/47542 (336161) | -0.07 | -0.08 | -0.06 | 1.34E$^{-39}$ |
| Pack years of smoking | 100944 | 0.04 | 0.03 | 0.05 | 3.13E$^{-39}$ |
| Cereal intake | 322455 | -0.04 | -0.05 | -0.04 | 4.03E$^{-39}$ |
| Illness, injury, bereavement, stress in last 2 years: Financial difficulties | 297736/37257 (334993) | 0.07 | 0.06 | 0.08 | 5.05E$^{-38}$ |
| Average total household income before tax | 290450 | -0.04 | -0.05 | -0.04 | 3.79E$^{-37}$ |
| Arm predicted mass (right) | 330982 | 0.01 | 0.01 | 0.02 | 6.46E$^{-37}$ |

| | | | | | |
|---|---|---|---|---|---|
| Pack years adult smoking as proportion of life span exposed to smoking | 100944 | 0.04 | 0.03 | 0.05 | 9.04E-37 |
| Basal metabolic rate | 331076 | 0.01 | 0.01 | 0.02 | 1.19E-36 |
| Attendance/disability/mobility allowance: Disability living allowance | 320532/14330 (334862) | 0.11 | 0.09 | 0.12 | 2.10E-36 |
| Seen doctor (GP) for nerves, anxiety, tension or depression | 219601/115287 (334888) | 0.05 | 0.04 | 0.05 | 9.26E-36 |
| Arm fat-free mass (right) | 330988 | 0.01 | 0.01 | 0.02 | 1.12E-35 |
| Age started smoking in former smokers | 81817 | -0.04 | -0.05 | -0.04 | 5.86E-35 |
| Age started oral contraceptive pill | 144192 | -0.03 | -0.03 | -0.02 | 1.47E-34 |
| Impedance of arm (left) | 331060 | -0.01 | -0.02 | -0.01 | 9.67E-34 |
| Qualifications: O levels/GCSEs or equivalent | 173754/160105 (333859) | -0.04 | -0.05 | -0.04 | 3.63E-33 |
| Number of treatments/medications taken | 336947 | 0.04 | 0.03 | 0.04 | 7.66E-33 |
| Father's age at death | 248380 | -0.02 | -0.03 | -0.02 | 8.45E-33 |
| Time spend outdoors in summer | 306336 | 0.04 | 0.03 | 0.05 | 1.01E-32 |
| Year ended full time education | 86337 | -0.02 | -0.02 | -0.01 | 2.16E-32 |
| Gamma glutamyltransferase | 321101 | 0.02 | 0.02 | 0.02 | 7.38E-32 |
| Risk taking | 242684/82908 (325592) | 0.05 | 0.04 | 0.06 | 2.38E-31 |
| Current employment status: Unable to work because of sickness or disability | 323591/12458 (336049) | 0.11 | 0.09 | 0.12 | 4.53E-31 |
| Time spent watching television (TV) | 319553 | 0.04 | 0.03 | 0.05 | 5.04E-31 |
| Amount of alcohol drunk on a typical drinking day | 101397 | 0.07 | 0.06 | 0.08 | 6.00E-31 |
| Frequency of consuming six or more units of alcohol | 101607 | 0.07 | 0.06 | 0.08 | 9.76E-31 |
| Liking for cabbage | 127891 | 0.06 | 0.05 | 0.07 | 1.21E-30 |
| Weekly usage of mobile phone in last 3 months | 281062 | 0.04 | 0.03 | 0.05 | 1.88E-30 |
| White blood cell (leukocyte) count | 326996 | 0.02 | 0.02 | 0.02 | 3.65E-30 |
| Leg fat-free mass (left) | 331026 | 0.01 | 0.01 | 0.01 | 2.06E-29 |
| Illnesses of mother: Chronic bronchitis/emphysema | 291513/18127 (309640) | 0.09 | 0.07 | 0.10 | 2.65E-29 |
| Leg predicted mass (left) | 331022 | 0.01 | 0.01 | 0.01 | 4.26E-29 |
| Impedance of arm (right) | 331049 | -0.01 | -0.02 | -0.01 | 1.01E-28 |

| | | | | | |
|---|---|---|---|---|---|
| Patient classification on admission (recoded): Inpatient | 132820/204168 (336988) | 0.04 | 0.03 | 0.05 | 1.57E$^{-28}$ |
| Whole body fat-free mass | 331060 | 0.01 | 0.01 | 0.01 | 1.56E$^{-27}$ |
| Ever taken cannabis | 110342 | 0.08 | 0.07 | 0.10 | 1.72E$^{-27}$ |
| Ease of skin tanning | 330266 | -0.03 | -0.04 | -0.03 | 1.72E$^{-27}$ |
| Whole body water mass | 331086 | 0.01 | 0.01 | 0.01 | 3.02E$^{-27}$ |
| Types of physical activity in last 4 weeks | 315948/19431 (335379) | 0.08 | 0.07 | 0.09 | 5.71E$^{-27}$ |
| Taking other prescription medications | 178431/157682 (336113) | 0.04 | 0.03 | 0.04 | 5.80E$^{-27}$ |
| Mother's age | 130898 | -0.02 | -0.02 | -0.01 | 6.22E$^{-27}$ |
| Qualifications: Other professional qualifications eg: nursing, teaching | 235971/97888 (333859) | -0.04 | -0.05 | -0.03 | 1.10E$^{-26}$ |
| Alcohol intake versus 10 years previously | 313088 | 0.04 | 0.03 | 0.04 | 1.19E$^{-26}$ |
| Pain type(s) experienced in last month: Back pain | 251368/85080 (336448) | 0.04 | 0.03 | 0.05 | 3.37E$^{-26}$ |
| Liking for gherkins | 125076 | 0.05 | 0.04 | 0.06 | 4.48E$^{-26}$ |
| Mean sphered cell volume | 321653 | 0.02 | 0.02 | 0.02 | 6.13E$^{-26}$ |
| Miserableness | 189898/141688 (331586) | 0.04 | 0.03 | 0.04 | 7.57E$^{-26}$ |
| Liking for cigarette smoking | 118235 | 0.12 | 0.09 | 0.14 | 7.86E$^{-26}$ |
| Fed-up feelings | 196440/133906 (330346) | 0.04 | 0.03 | 0.04 | 8.73E$^{-26}$ |
| Operative procedures - secondary OPCS4: Z94.3 Left sided operation | 232690/104298 (336988) | 0.04 | 0.03 | 0.05 | 1.65E$^{-25}$ |
| Operative procedures - OPCS4: Z94.3 Left sided operation | 232690/104298 (336988) | 0.04 | 0.03 | 0.05 | 1.65E$^{-25}$ |

**Supplementary Table S2. Details of plausible phenotypes in UK Biobank**

| Phenotype | Biobank field number | Phenotype description |
|---|---|---|
| Body mass index (BMI) | 21001 | Calculated from measures of standing height (using a Seca 202 device, cm) and weight (measured by a variety of means, Kg) collected during visits (Kg/m2). |
| Body fat percentage | 23099 | Body composition estimation by impedance measurement, as a percentage in 0.1% increments. Measured using the Tanita BC418MA body composition analyser. |
| Wheeze or whistling in the chest in last year | 2316 | Participants asked* "In the last year have you ever had wheeze or whistling in the chest?", (yes/no). |
| C-reactive protein[1] | 30710 | Measured by immunoturbidimetric – high sensitivity analysis on a Beckman Coulter AU5800 (mg/L). |
| Date first reported (other chronic obstructive pulmonary disease) | 131492 | Date of the first occurrence of any code mapped to the 3-character ICD10 J44. This code corresponds to "other chronic obstructive pulmonary disease". |
| Mouth/teeth dental problems: Dentures | 6149#6 | Participants asked* "Do you have any of the following?", of which dentures was one of the options. We created a binary (yes/no) variable from this for dentures. |
| Overall health rating | 2178 | Participants asked* "In general how would you rate your overall health?". The options were: excellent, good, fair and poor. In our results we refer to this as poorer health rating due to the coding being in this direction. |
| Gamma glutamyl transferase[1] | 30730 | Measured by enzymatic rate method on a Beckman Coulter AU5800 (U/L). |
| White blood cell (leukocyte) count[1] | 30000 | Result of white blood cell count, performed on blood samples obtained at visits, i.e., number of leukocytes. The Beckman analyser was used, and samples were typically analysed within 24 hours of blood draw ($10^9$ cells/L). |
| Mean sphered cell volume | 30270 | Obtained from the Beckman Coulter LH750 (femtolitres). |
| Seen doctor (GP) for nerves, anxiety, tension or depression | 2090 | Participants asked* "Have you ever seen a general practitioner (GP) for nerves, anxiety, tension or depression?", (yes/no). |
| Number of treatments/ medications taken[1] | 137 | The number of treatments (medications) entered (as a count of field 20003). This was from a verbal interview by a trained nurse on prescription medications. If the participant responded that they were taking regular medication, then they were asked if they could tell the |

| | | |
|---|---|---|
| | | interviewer what these were. This included medication taken regularly and not short-term medications, prescribed medication not taken, over-thx10-counter medications or vitamins and supplements. |
| Amount of alcohol drunk on a typical drinking day | 20403 | Participants asked* “How many drinks containing alcohol do you have on a typical day when you are drinking?”, where a ‘drink’ was defined as one unit of alcohol. Options were 1 or 2, 3 or 4, 5 or 6, 7,8 or 9 and 10 or more. Only participants who had indicated they drink alcohol in the question “how often do you have a drink containing alcohol?”. |

* Participants were asked these questions as part of the Assessment Centre Environment (ACE) touch screen questionnaire. Participants could also answer “do not know” or “prefer not to answer” or similar for most questions, but these data were removed for analyses.

[1]Variables that were inverse normal rank transformed

**Supplementary Table S3. Details of implausible phenotypes in UK Biobank**

| Phenotype | Biobank ID | Phenotype description |
|---|---|---|
| Lifetime number of sexual partners[1] | 2149 | Participants asked* "About how many sexual partners have you had in your lifetime?". Only asked to those participants who had indicated that they had previously had sexual intercourse. Responses <1 or >99997 were rejected and participants were asked to confirm if response was >99. |
| Age at first live birth[2] | 2754 | Participants asked* "How old were you when you had your first child?". Only asked to those participants who indicated they had given birth to more than one child. Responses <8, >65, >participants age or <age at first period were rejected and participants asked to confirm if response was <12, >48, >age when periods stopped. |
| Townsend deprivation index at recruitment | 189 | Calculated immediately prior to joining UK Biobank, based on the national census output area that the participant resides in. A higher score indicates greater levels of socioeconomic deprivation. |
| Leisure/social activities: Religious group | 6160 (3) | Participants asked* "Which of the following do you attend once a week or more often?". Participants could select multiple options of which religious group was one of the options. We created a binary (yes/no) variable from this for religious group. |
| Cereal intake[1] | 1458 | Participants asked* "How many bowls of cereal do you eat a week?". Responses <0 or >99 were rejected, and participants asked to confirm if response was >14. For those participants that selected less than one bowl per week, we assigned them to the value 1 bowls per week, therefore a value of 1 actually represents 0-1. |
| Risk taking | 2040 | Participants asked* "Would you describe yourself as someone who takes risks?", (yes/no). |
| Time spent watching television[1] | 1070 | Participants asked* "In a typical day, how many hours do you spend watching TV? (Put 0 if you do not spend any time doing it?)". Responses <0 or >24 were rejected, and participants asked to confirm if response was >8. For those participants that selected less than one hour per day, we assigned them to the value 1 hour per day, therefore a value of 1 actually represents 0-1. |
| Liking for cabbage | 20630 | This question was part of an online follow-up study about food and other preferences, where they were asked about liking for cabbage on a 9-point scale. Only values 1 (extremely dislike), 5 (neither like nor dislike) and 9 (extremely like) were assigned explicit meanings with other values lying between these. Participants could also indicate they had never tried to item. |
| Weekly usage of mobile | 1120 | Participants asked* "Over the last 3 months, on average how much time per week did you spend making or receiving calls on |

| | | |
|---|---|---|
| phone in last 3 months | | a mobile phone?". This was only asked to participants who had indicated they used a mobile phone at least once per week in the past or did not know whether they had used it. The options were: less than 5 mins, 5-29 mins, 30-59 mins, 1-3 hours, 4-6 hours and more than 6 hours. |
| Ease of skin tanning | 1727 | Participants asked* "What would happen to your skin if it was repeatedly exposed to bright sunlight without any protection. The options were: get very tanned, get moderately tanned, get mildly or occasionally tanned, never tan, only burn. |
| Mother's age at time of questionnaire | 1845 | Participants asked* "what is her age now?". This was only asked to participants who had indicated that their mother (or adopted mother) was still alive. Responses <participants age + 10 years or >122 were rejected, and participants asked to confirm if response was greater than participant's age by 14 years or >105. |
| Pain type(s) experienced in last month (back pain) | 6159 (4) | Participants asked* "In the last month have you experienced any of the following that interfered with your usual activities? (You can select more than one answer)?". Participants could select multiple options of which back pain was one of the options. We created a binary (yes/no) variable from this for back pain. |
| Had an operation on the left-side of the body | 41210 (Z943) | Obtained from hospital inpatient records. Operative procedures are coded according to the office of Population Censuses and Surveys Classification of Interventions and Procedures, version 4 (OPCS-4). Z943 is indicative of a left sided operation and we created a binary (yes/no) variable from this. |

* Participants were asked these questions as part of the Assessment Centre Environment (ACE) touch screen questionnaire. Participants could also answer "do not know" or "prefer not to answer" or similar for most questions, but these data were removed for analyses.

[1]Variables that were inverse normal rank transformed

[2]Opposite to any adverse effect on fertility in phenome-wide association study

**Supplementary Table S4. ALSPAC phenotypes for plausible phenotypes**

| Phenotype | Sample | ALSPAC questionnaire or clinic timepoint | Phenotype description |
|---|---|---|---|
| Body mass index (BMI) | Mother | FOM clinic | Calculated from measures of height (using a Harpenden stadiometer to the nearest 1mm, m) and weight (using the Tanita scales to the nearest 0.1Kg, Kg) collected during visits ($Kg/m^2$). |
| | Father | FOF clinic | Calculated from measures of height (using a Harpenden stadiometer to the nearest 1mm, m) and weight (using the Tanita scales to the nearest 0.1Kg, Kg) collected during visits ($Kg/m^2$). |
| | Child | Age 7 clinic | Calculated from measures of height (using a Harpenden stadiometer to the nearest 1mm, cm) and weight (using the Tanita body fat analyser to the nearest 50g, Kg) collected during visits ($Kg/m^2$). |
| Body fat percentage | Mother | FOM clinic | Calculated from body fat mass (g) and weight (Kg, described above). Body fat mass was measured using Dual Emission X-ray Absorptiometry (DXA, Lunar Prodigy). This measure was obtained from a full body scan (not obtained in pregnant women). |
| | Father | FOF clinic | Calculated from body fat mass (g) and weight (Kg, described above). Body fat mass was measured using Dual Emission X-ray Absorptiometry (DXA, Lunar Prodigy). This measure was obtained from a full body scan. |
| | Child | Age 9 clinic | Calculated from body fat mass (g) and weight (Kg, described above but used weight collected at age 9 clinic here). Body fat mass was measured using Dual Emission X-ray Absorptiometry (DXA, Lunar Prodigy). This measure was obtained from a full body scan. |
| Wheeze or whistling in the chest | Mother | Pregnancy | Participants asked "Have you had any of the following in the past two years: attacks of wheezing with whistling on the chest". The options were: yes (often or sometimes); no. |
| | Father | Pregnancy | Participants asked "Have you had any of the following in the past two years: attacks of wheezing with whistling on the chest?". The options were: yes (often or sometimes); no. |

| | | | |
|---|---|---|---|
| | Child | Child aged 6 months (T1) and 30 (T2) months | T1: Mother was asked “Has your baby had any of the following: wheezing?”. The options were: yes (often or sometimes); no.<br><br>T2: Mother was asked “Has he/she had any of the following since he/she was 18 months old: wheezing?”. The options were: yes (often or sometimes); no. |
| C-reactive protein (CRP) | Mother | FOM clinic[1] | Obtained from fasting blood samples which, after collection, were spun, frozen and stored at -80$^{o}$C. High-sensitivity CRP concentrations were measured using an automated particle-enhanced immunoturbidimetric assay (Roche, Welwyn Garden City, UK) with a minimum detection limit of 0.01 mg/L. (mg/L) |
| | Father | FOF clinic[1] | Obtained from fasting blood samples which, after collection, were spun, frozen and stored at -80$^{o}$C. High-sensitivity CRP concentrations were measured using an automated particle-enhanced immunoturbidimetric assay (Roche, Welwyn Garden City, UK) with a minimum detection limit of 0.01 mg/L. (mg/L) |
| | Child | Age 9 clinic[1] | Obtained from non-fasting blood samples which, after collection, were spun, frozen and stored at -80$^{o}$C. High-sensitivity CRP concentrations were measured using an automated particle-enhanced immunoturbidimetric assay (Roche, Welwyn Garden City, UK) with a minimum detection limit of 0.01 mg/L. (mg/L) |
| Overall health rating | Mother | Child aged 8 weeks (T1) and 8 months (T2) | T1: Participants asked “How would you describe your health now?”. The options were: hardly ever well; often unwell; mostly well; always well.<br><br>T2: Participants asked “Which of the following would you say describes your health now?”. The options were: hardly ever feel really well; often feel unwell; mostly feel well and healthy; always fit and well. |
| | Father | Child aged 8 weeks (T1) and 8 months (T2) | T1: Participants asked “How would you describe your health now?”. The options were: hardly ever well; often unwell; mostly fit and well; always fit and well.<br><br>T2: Participants asked “Which of the following would you say describes your health now?”. |

| | | | |
|---|---|---|---|
| | | | The options were: hardly ever feel really well; often feel unwell; mostly feel well and healthy; always fit and well. |
| | Child | Child aged 4 weeks (T1) and 6 months (T2) | T1: Participants asked “How would you describe the health of your baby now?”. The options were: almost always unwell; sometimes quite ill; healthy but a few minor problems; very healthy.<br><br>T2: Participants asked “How would you assess the health of your baby?”. The options were: almost always unwell; sometimes quite ill; healthy but a few minor problems; very healthy no problems. |
| Gamma glutamyl transferase | Child | Age 9 clinic | Obtained from non-fasting blood samples which, after collection, were spun, frozen and stored at -80$^{o}$C. High-sensitivity CRP concentrations were measured using an automated particle-enhanced immunoturbidimetric assay (Roche, Welwyn Garden City, UK) with a minimum detection limit of 0.01 mg/L. (U/L) |
| Seen doctor (GP) for anxiety/nerves or depression (separate phenotypes) | Mother | Child aged 8 months (T1) and 21 months (T2) | T1: Participants asked “Have you had any of the following since the baby was born: a) anxiety or nerves, b) depression”. The options were: yes and saw doctor; yes did not see doctor; no.<br><br>T2: Participants asked “Have you had any of the following since your toddler was 8 months old: a) anxiety or nerves, b) depression”. The options were: yes and saw doctor; yes did not see doctor; no. |
| | Father | Child aged 8 months (T1) and 21 months (T2) | T1: Participants asked “Have you had any of the following since the new baby was born: a) anxiety or nerves, b) depression”. The options were: yes and consulted doctor, yes but did not consult doctor, no.<br><br>T2: Participants asked “Have you had any of the following since your toddler was 8 months old: a) anxiety or nerves, b) depression”. The options were: yes and consulted doctor; yes but did not consult doctor; no. |
| | Child | Child aged 91 months | The number of general anxiety symptoms from the following items for which “yes, but not on most days” or “yes happened on more |

| | | | |
|---|---|---|---|
| | | | days than not" were ticked (other options was "no not at all") was calculated and an indicator for whether any general anxiety symptoms were reported was created.<br><br>Questions:<br><br>a) Does worrying lead to him/her being restless, feeling keyed up, tense or on edge, or being unable to relax?<br><br>b) Does worrying lead to him/her feeling tired or 'worn out' more easily?<br><br>c) Does worrying lead to difficulties on concentrating or his/her mind going blank?<br><br>d) Does worrying lead to irritability?<br><br>e) Does worrying lead to him/her looking physically tense (tense muscles)?<br><br>f) Does worrying interfere with his/her sleep (e.g., difficulty in falling or staying asleep, or restless sleep, or doesn't have a good night's sleep)? |
| Number of medications taken | Mother | Child aged 8 weeks (T1)[1] and 21 months (T2)[1] | T1: The number of medications was calculated from the following question: "Please name all the pills, medicines or ointments you are currently using or have used since the baby was born."<br><br>T2: The number of medications was calculated from the following question: "Please list all the medicines and pills that you have taken in the past month." |
| | Father | Child aged 21 months (T1)[1] and 73 months (T2)[1] | T1: The number of medications was calculated from the following question: "Please name all the medicines, pills and ointments that you have taken in the past month."<br><br>T2: The number of medications was calculated from the following question: "Please list all the drugs, medicines and ointments that you have taken in the past month." |
| | Child | Child aged 4 weeks (T1)[1] | T1: The number of medications was calculated from the following question: "Please list all the |

| | | | |
|---|---|---|---|
| | | and 24 months (T2) | ointments, pills and medicines that have been given to your baby while he/she has been at home.”<br><br>T2: The number of different types of medications was calculated from the following question: “Children often have accidents or illnesses that need treatment. Please indicate which of the following have been given to your child since he was 15 months old”. The options were: cough medicine; antibiotics/penicillin; throat medicine; vitamins; paracetamol/calpol; ointment for skin; eye ointment; diarrhoea mixture or pills; dimotapp/decongestant; ear drops; eye drops; teething gel; laxative; other (please describe). |
| Amount of alcohol drunk | Mother | Pregnancy | Participants asked “How often have you drunk alcoholic drinks? Please indicate for each of the following times: Before this pregnancy?”. The options were: never; less than 1 glass a week; at least 1 glass a week; 1-2 glasses every day; at least 3-9 glasses every day; at least 10 glasses every day” |
| | Father | Pregnancy | Participants asked “How often have you drunk alcoholic drinks: Before your partner became pregnant?”. The options were: never; less than once a week; at least once a week; 1-2 glasses every day,; 3-9 glasses every day; at least 10 glasses every day” |

[1]Variables that were inverse normal rank transformed. FOM = focus on mothers, FOF = focus on fathers, T1 = time point 1, T2 = time point 2

**Supplementary Table S5. ALSPAC phenotypes for implausible phenotypes**

| Phenotype | Sample | ALSPAC questionnaire or clinic timepoint | Phenotype description |
|---|---|---|---|
| Age at first pregnancy | Mother | Pregnancy | Participant asked “How old were you when you became pregnant for the very first time?” |
| Townsend deprivation index at recruitment | Mother | Pregnancy (T1) and child aged 8 weeks (T2) | Postcode data for participants has been linked to publicly available data on Townsend deprivation scores. Each variable is coded as quintiles (5 categories) to minimise disclosure risk. This data is available for all mother questionnaires, and we have used T1 and T2 in this study. A higher score indicates greater levels of socioeconomic deprivation. |
| Attends a place of worship | Mother | Pregnancy | Participant asked “Do you go to a place of worship?”. The options were: yes, at least once a week; yes, at least once a month; yes, at least once a year; not at all. |
| | Father | Pregnancy | Participant asked “Do you go to a place of worship?”. The options were: yes, at least once a week; yes, at least once a month; yes, at least once a year; not at all. |
| | Child | Child aged 115 months | Participant asked “Does he/she attend a place of worship (church, mosque, etc)?”. The options were: yes, often; yes, sometimes; no, not at all. |
| Cereal intake (separate phenotypes for oat, wholegrain or bran and other) | Mother | Pregnancy (32 weeks gestation) (T1) and child aged 47 months (T2) | T1: Participants asked: “How many times a week nowadays do you eat:<br><br>l) Oat cereals (e.g., porridge, Ready Brek, Muesli)<br><br>m) Wholegrain or bran cereals (e.g., All bran, Bran flakes, Weetabix, Wheatflakes, Fruit and Fibre)<br><br>n) Other cereals (e.g., Cornflakes, Rice Krispies, Special K, Frosties)”<br><br>The options were: Never or rarely; once in 2 weeks; 1 to 3 times per week; 4 to 7 times per week; more than once a day. |

| | | | |
|---|---|---|---|
| | | | T2: Participants asked: “Mothers eat a variety of different things. How often nowadays do you eat the following foods? Please answer every question even if you never eat the food (in this case tick ‘Never or rarely’:<br><br>l) Oat cereals (e.g., porridge, Ready Brek, Muesli)<br><br>m) Wholegrain or bran cereals (e.g., All bran, Bran flakes, Weetabix, Wheatflakes, Fruit and Fibre, Shredded Wheat)<br><br>n) Other cereals (e.g., Cornflakes, Rice Krispies, Special K, Frosties)”<br><br>The options were: Never or rarely; once in 2 weeks; 1 to 3 times per week; 4 to 7 times per week; more than once a day. |
| | Father | Child aged 47 months | Participants asked: “How often nowadays do you eat the following foods. Please answer every question even if you never eat the food (in this case tick ‘Never or rarely’:<br><br>l) Oat cereals (e.g., porridge, Ready Brek, Muesli)<br><br>m) Wholegrain or bran cereals (e.g., All bran, Bran flakes, Weetabix, Wheatflakes, Fruit and Fibre, Shredded wheat)<br><br>n) Other cereals (e.g., Cornflakes, Rice Krispies, Special K, Frosties)”<br><br>The options were: Never or rarely; Once in 2 weeks; 1 to 3 times per week; 4 to 7 times per week; more than once a day. |
| | Child | Child aged 54 months | After being asked “Does she/he eat breakfast cereals at all?”. Participants who answered yes, were then asked: “What type of breakfast cereal does she/he eat nowadays:<br><br>l) Oat cereals (e.g., porridge, Ready Brek, Muesli, chocolate Ready Brek) |

| | | | |
|---|---|---|---|
| | | | m) Wholegrain or bran cereals (e.g., All bran, Bran flakes, Weetabix, Wheatflakes, Fruit and Fibre, Shreddies, Shredded wheat, Sugar puffs)<br><br>n) Other cereals (e.g., Cornflakes, Rice Krispies, Frosties, Special K, Coco pops)"<br><br>The options were: Never or rarely; Once in 2 weeks; 1 to 3 times per week; 4 to 7 times per week; more than once a day. |
| Risk taking | Child | Child aged 30 months (T1) and 42 months (T2) | T1: Participants were asked "How often does he/she avoid taking risks?". The options were: never; hardly ever; sometimes; often; very often.<br><br>T2: Participants were asked "How often does he/she avoid taking risks?". The options were: never; hardly ever; sometimes; often; very often. |
| Time spent watching television (separate variables for weekend day and weekday) | Mother | Child aged 85 months[1] | Participants were asked "On average how many hours per day do you spend doing the following: watching television". Participants could respond for weekdays and weekend days.<br><br>Any values above 16 were removed as this is likely a mistake in reading the question. |
| | Father | Child aged 85 months[1] | Participants were asked "On average how many hours per day do you spend doing the following: watching television". Participants could respond for weekdays and weekend days.<br><br>Any values above 16 were removed as this is likely a mistake in reading the question. |
| | Child | Child aged 38 months | Participants were asked "How much time on average does she/he spend on most weekdays watching T.V" and "How much time on average does she/he spend on most weekend days watching T.V". The options for both were: not at all; less than 1 hour; 1 to 2 hours a day; more than 2 hours a day. |
| Cabbage intake | Mother | Pregnancy (32 weeks gestation) (T1) and child | T1: Participants asked: "How many times a week nowadays do you eat: Cabbage, brussel sprouts, kale and other green leafy vegetables?" |

| | | | |
|---|---|---|---|
| | | aged 47 months (T2) | The options were: Never or rarely; once in 2 weeks; 1 to 3 times per week; 4 to 7 times per week; more than once a day.<br><br>T2: Participants asked: “How many times nowadays do you eat: Cabbage, brussel sprouts, kale and other green leafy vegetables?”<br><br>The options were: Never or rarely; Once in 2 weeks; 1 to 3 times per week; 4 to 7 times per week; more than once a day. |
| | Father | Pregnancy | Participants asked: “We are interested in your diet - how many times each week nowadays do you eat: Cabbage, brussel sprouts, kale and other green leafy vegetables?”<br><br>The options were: Never or rarely; Once in 2 weeks; 1 to 3 times per week; 4 to 7 times per week; more than once a day. |
| | Child | Child aged 38 months (T1) and 54 months (T2) | T1: Participants asked: “How many times nowadays does she/he eat: Cabbage, brussel sprouts, kale and other green leafy vegetables?”<br><br>The options were: Never or rarely; Once in 2 weeks; 1 to 3 times per week; 4 to 7 times per week; more than once a day.<br><br>T2: Participants asked: “How many times nowadays does she/he eat: Cabbage, brussel sprouts, kale and other green leafy vegetables?”<br><br>The options were: Never or rarely; Once in 2 weeks; 1 to 3 times per week; 4 to 7 times per week; more than once a day. |
| Mobile phone usage | Mother | Child aged 85 months | Participants were first asked if they have a mobile phone and if they answered yes they were asked: “How often do you use it?”.<br><br>The options were: At least once a day; 4 to 6 times a week; 1 to 3 times a week; less than once a week. |

| | | | |
|---|---|---|---|
| | Father | Child aged 85 months | Participants were first asked if they have a mobile phone and if they answered yes they were asked: “How often do you use it?”.<br><br>The options were: At least once a day; 4 to 6 times a week; 1 to 3 times a week; less than once a week. |
| | Child | Child aged 122 months | Participants asked: “How long altogether do you usually use a mobile phone now?” They were asked to respond for “on a school day” and “on a weekend day”.<br><br>The options were: Not at all; less than 15 minutes; 15 to 30 minutes; more than 30 minutes. |
| Mother’s age at time of questionnaire | Mother | Pregnancy (32 weeks gestation) | This was calculated from the age of the participant when the baby was born and her own mothers age at the time she was born. For the latter participants were asked: “How old was your natural mother when you were born?”. |
| | Father | Pregnancy | This was calculated from the age of the participant and their own mothers age at the time they were born. For the latter participants were asked: “How old was your mother when you were born?”. |
| Back pain | Mother | Child aged 8 weeks (T1) and 8 months (T2) | T1: Participants asked: “Since having the baby have the following occurred: backache?”<br><br>The options were: yes (almost always or sometimes); not at all.<br><br>T2: Participants asked: “Have you had any of the following since the baby was born: backache?”<br><br>The options were: yes; no. |
| | Father | Pregnancy | Participants asked: “Have you ever had any of the following problems:<br><br>The options were: yes (recently or in the past), no never. |

[1]Variables that were inverse normal rank transformed. T1 = time point 1, T2 = time point 2

**Supplementary Table S6. Summary statistics for UK Biobank sample with data available**

| Phenotype | N | Mean (SD) or percentage (%) |
|---|---|---|
| Ever smoked | 335,804 | 45.20% (9.93% current and 35.27% former) |
| Smoking heaviness (CPD, for current or former smokers) | 100,150 | 18.25 (10.10) |
| Lifetime smoking score | 335,820 | 0.34 (0.68) |
| Age (years) | 336,988 | 56.87 (8.00) |
| Sex (females) | 336,988 | 53.79% |
| Body mass index (Kg/m$^2$) | 335,908 | 27.39 (4.75) |
| Body fat percentage | 330,893 | 31.35 (8.52) |
| Wheeze or whistling in the chest in last year | 331,124 | 20.55% |
| C-reactive protein (mg/L) | 320,589 | 2.58 (4.37) |
| Ever reported COPD | 336,988 | 3.11% |
| Had dentures | 335,919 | 12.95% |
| Overall health rating (percentage in each group) | 335,806 | Excellent = 16.87%<br>Good = 58.68%<br>Fair = 20.32%<br>Poor = 4.12% |
| Gamma glutamyl transferase (U/L) | 321,101 | 37.30 (41.73) |
| White blood cell count ($10^9$ cells/L) | 326,996 | 6.89 (2.01) |
| Mean sphered cell volume (femtolitres) | 321,653 | 82.87 (5.25) |
| Seen GP for nerves, anxiety, tension or depression | 334,888 | 34.42% |
| Number of treatments/ medications taken | 336,947 | 2.45 (2.65) |
| Alcohol drunk on typical drinking day (percentage in each group) | 101,397 | 1 or 2 = 51.07%<br>3 or 4 = 27.49%<br>5 or 6 = 12.10%<br>7,8 or 9 = 6.52%<br>10 or more = 2.83% |
| Lifetime number of sexual partners | 277,634 | 7.29 (64.03) |
| Age at first live birth | 124,051 | 25.39 (4.54) |
| Townsend deprivation index (higher score = more deprived) | 336,591 | -1.58 (2.93) |
| Takes part in a religious group | 336,161 | 8.59% |
| Cereal intake (bowls/week) | 336,304 | 4.66 (2.74) |
| Risk-taking (responded yes) | 325,592 | 25.46% |

| | | |
|---|---|---|
| Time spent watching television (hours/day) | 334,630 | 2.83 (1.61) |
| Liking for cabbage (percentage in each group) | 127,891 | 1 (extremely dislike) = 2.31%<br>2 = 1.47%<br>3 = 2.10%<br>4 = 3.15%<br>5 (neither like nor dislike) = 7.49%<br>6 = 13.26%<br>7 = 23.17%<br>8 = 21.57%<br>9 (extremely like) = 25.48% |
| Weekly usage of mobile phone (percentage in each group) | 281,062 | Less than 5 mins = 21.81%<br>5-29 mins = 39.49%<br>30-59 mins = 17.00%<br>1-3 hours. = 13.72%<br>4-6 hours = 3.95%<br>More than 6 hours = 4.04% |
| Ease of skin tanning (percentage in each group, higher = less likely to tan) | 330,266 | Get very tanned = 20.11%<br>Get moderately tanned = 40.78%<br>Get mildly or occasionally tanned = 21.54%<br>Never tan, only burn = 17.58% |
| Mother's age at time of questionnaire | 130,898 | 78.59 (8.10) |
| Back pain experienced in last month | 336,448 | 11.89% |
| Operation on the left-side of the body | 336,988 | 1.53% |

[1]Lifetime smoking score is standardised, CPD=cigarettes per day, COPD=chronic obstructive pulmonary disease

**Supplementary Table S7. Associations between the polygenic risk scores for lifetime smoking score, smoking heaviness and smoking initiation and plausible outcomes in UK Biobank**

| Outcome | Model | N | Beta or OR (95% CI) | P-value |
|---|---|---|---|---|
| ***Lifetime smoking score PRS*** | | | | |
| BMI | Linear | 335,908 | 0.09 (0.08 to 0.11) | $1.06 \times 10^{-29}$ |
| Body fat percentage | Linear | 330,893 | 0.11 (0.09 to 0.14) | $1.66 \times 10^{-25}$ |
| Wheeze | Logistic | 331,124 | 1.04 (1.03 to 1.05) | $1.21 \times 10^{-17}$ |
| C-reactive protein | Linear | 320,589 | 0.02 (0.01 to 0.02) | $1.17 \times 10^{-21}$ |
| Ever reported COPD | Logistic | 336,988 | 1.11 (1.08 to 1.13) | $2.10 \times 10^{-23}$ |
| Had dentures | Logistic | 335,919 | 1.04 (1.03 to 1.05) | $2.06 \times 10^{-12}$ |
| Poorer health rating | Ordinal | 335,806 | 1.03 (1.02 to 1.04) | $6.24 \times 10^{-19}$ |
| Gamma glutamyl transferase | Linear | 321,101 | 0.01 (0.01 to 0.02) | $4.50 \times 10^{-19}$ |
| White blood cell count | Linear | 326,996 | 0.01 (0.01 to 0.02) | $1.93 \times 10^{-16}$ |
| Mean sphered cell volume | Linear | 321,653 | 0.03 (0.01 to 0.05) | $2.77 \times 10^{-03}$ |
| Seen GP for nerves, anxiety, tension or depression | Logistic | 334,888 | 1.02 (1.01 to 1.02) | $6.60 \times 10^{-06}$ |
| Number of treatments/ medications taken | Linear | 336,947 | 0.01 (0.01 to 0.01) | $4.86 \times 10^{-12}$ |
| Alcohol drunk on typical drinking day | Ordinal | 101,397 | 1.03 (1.02 to 1.04) | $6.24 \times 10^{-06}$ |
| ***Smoking heaviness PRS – never smokers*** | | | | |
| BMI | Linear | 183,478 | 0.02 (-0.002 to 0.04) | 0.07 |
| Body fat percentage | Linear | 180,874 | 0.02 (-0.01 to 0.04) | 0.29 |
| Wheeze | Logistic | 181,268 | 1.003 (0.99 to 1.02) | 0.58 |
| C-reactive protein | Linear | 175,026 | 0.002 (-0.002 to 0.007) | 0.29 |
| Ever reported COPD | Logistic | 184,005 | 1.00 (0.95 to 1.05) | 0.96 |
| Had dentures | Logistic | 183,439 | 1.00 (0.98 to 1.02) | 0.93 |
| Poorer health rating | Ordinal | 183,522 | 1.002 (0.99 to 1.01) | 0.74 |
| Gamma glutamyl transferase | Linear | 175,282 | 0.00007 (-0.004 to 0.004) | 0.98 |
| White blood cell count | Linear | 178,443 | -0.003 (-0.007 to 0.002) | 0.26 |
| Mean sphered cell volume | Linear | 175,491 | -0.01 (-0.04 to 0.01) | 0.30 |
| Seen GP for nerves, anxiety, tension or depression | Logistic | 182,924 | 1.00 (0.99 to 1.01) | 0.51 |
| Number of treatments/ medications taken | Linear | 183,980 | 0.004 (-0.0003 to 0.008) | 0.07 |
| Alcohol drunk on typical drinking day | Ordinal | 58,040 | 1.00 (0.98 to 1.02) | 0.98 |
| ***Smoking heaviness PRS – former smokers*** | | | | |
| BMI | Linear | 118,082 | 0.02 (-0.007 to 0.05) | 0.15 |
| Body fat percentage | Linear | 116,254 | 0.01 (-0.03 to 0.04) | 0.58 |

| | | | | |
|---|---|---|---|---|
| Wheeze | Logistic | 116,544 | 1.01 (1.00 to 1.03) | 0.08 |
| C-reactive protein | Linear | 112,756 | 0.007 (0.001 to 0.01) | 0.02 |
| Ever reported COPD | Logistic | 118,447 | 1.04 (1.007 to 1.07) | 0.01 |
| Had dentures | Logistic | 118,128 | 1.01 (0.99 to 1.03) | 0.18 |
| Poorer health rating | Ordinal | 118,020 | 1.01 (1.00 to 1.03) | 0.02 |
| Gamma glutamyl transferase | Linear | 112,958 | -0.002 (-0.008 to 0.003) | 0.41 |
| White blood cell count | Linear | 115,058 | 0.004 (-0.001 to 0.01) | 0.12 |
| Mean sphered cell volume | Linear | 113,157 | -0.01 (-0.04 to 0.02) | 0.51 |
| Seen GP for nerves, anxiety, tension or depression | Logistic | 117,745 | 1.00 (0.99 to 1.02) | 0.64 |
| Number of treatments/ medications taken | Linear | 118,437 | 0.005 (-0.0002 to 0.01) | 0.06 |
| Alcohol drunk on typical drinking day | Ordinal | 92,224 | 0.99 (0.98 to 1.01) | 0.59 |
| ***Smoking heaviness PRS – current smokers*** | | | | |
| BMI | Linear | 33,177 | -0.10 (-0.15 to -0.05) | $2.19 \times 10^{-04}$ |
| Body fat percentage | Linear | 32,625 | -0.15 (-0.22 to -0.08) | $4.31 \times 10^{-05}$ |
| Wheeze | Logistic | 32,197 | 1.02 (0.99 to 1.04) | 0.15 |
| C-reactive protein | Linear | 31,694 | 0.01 (-0.0004 to 0.02) | 0.06 |
| Ever reported COPD | Logistic | 33,352 | 1.06 (1.03 to 1.10) | $5.06 \times 10^{-04}$ |
| Had dentures | Logistic | 33,208 | 0.98 (0.96 to 1.01) | 0.29 |
| Poorer health rating | Ordinal | 33,109 | 1.01 (0.99 to 1.03) | 0.48 |
| Gamma glutamyl transferase | Linear | 31,746 | -0.007 (-0.02 to 0.003) | 0.17 |
| White blood cell count | Linear | 32,360 | 0.006 (-0.005 to 0.02) | 0.27 |
| Mean sphered cell volume | Linear | 31,887 | 0.05 (-0.01 to 0.12) | 0.11 |
| Seen GP for nerves, anxiety, tension or depression | Logistic | 33,087 | 0.98 (0.96 to 1.00) | 0.06 |
| Number of treatments/ medications taken | Linear | 33,347 | -0.005 (-0.02 to 0.005) | 0.32 |
| Alcohol drunk on typical drinking day | Ordinal | 6,921 | 1.01 (0.97 to 1.05) | 0.68 |
| ***Smoking initiation PRS*** | | | | |
| BMI | Linear | 335,908 | 0.07 (0.06 to 0.09) | $1.82 \times 10^{-18}$ |
| Body fat percentage | Linear | 330,893 | 0.09 (0.07 to 0.11) | $3.76 \times 10^{-17}$ |
| Wheeze | Logistic | 331,124 | 1.03 (1.02 to 1.04) | $1.38 \times 10^{-13}$ |
| C-reactive protein | Linear | 320,589 | 0.01 (0.01 to 0.01) | $8.99 \times 10^{-08}$ |
| Ever reported COPD | Logistic | 336,988 | 1.07 (1.05 to 1.09) | $1.43 \times 10^{-10}$ |
| Had dentures | Logistic | 335,919 | 1.02 (1.01 to 1.03) | $4.48 \times 10^{-04}$ |
| Poorer health rating | Ordinal | 335,806 | 1.03 (1.02 to 1.03) | $1.40 \times 10^{-15}$ |
| Gamma glutamyl transferase | Linear | 321,101 | 0.01 (0.01 to 0.01) | $2.26 \times 10^{-09}$ |
| White blood cell count | Linear | 326,996 | 0.01 (0.01 to 0.01) | $1.45 \times 10^{-08}$ |
| Mean sphered cell volume | Linear | 321,653 | 0.04 (0.02 to 0.06) | $1.76 \times 10^{-05}$ |
| Seen GP for nerves, anxiety, tension or depression | Logistic | 334,888 | 1.03 (1.02 to 1.04) | $7.18 \times 10^{-14}$ |

| | | | | |
|---|---|---|---|---|
| Number of treatments/ medications taken | Linear | 336,947 | 0.01 (0.01 to 0.01) | $1.31x10^{-10}$ |
| Alcohol drunk on typical drinking day | Ordinal | 101,397 | 1.04 (1.03 to 1.05) | $1.85x10^{-11}$ |

For linear regression models the effect estimate is beta and for logistic and ordinal regressions this is the odds ratio. OR=odds ratio, CI=confidence interval, PRS=polygenic risk score, BMI=body mass index, COPD=chronic obstructive pulmonary disease, GP=general practitioner.

**Supplementary Table S8. Associations between the polygenic risk scores for lifetime smoking score, smoking heaviness and smoking initiation and implausible outcomes in UK Biobank**

| Outcome | Model | N | Beta or OR (95% CI) | P-value |
|---|---|---|---|---|
| ***Lifetime smoking score PRS*** | | | | |
| Lifetime number of sexual partners | Linear | 277,634 | 0.01 (0.01 to 0.02) | $4.64 \times 10^{-12}$ |
| Age at first live birth | Linear | 124,051 | -0.12 (-0.14 to -0.09) | $5.30 \times 10^{-20}$ |
| Townsend deprivation index | Linear | 336,591 | 0.04 (0.03 to 0.05) | $1.23 \times 10^{-17}$ |
| Takes part in a religious group | Logistic | 336,161 | 0.96 (0.95 to 0.98) | $8.44 \times 10^{-09}$ |
| Cereal intake | Linear | 336,304 | -0.01 (-0.02 to -0.01) | $1.10 \times 10^{-12}$ |
| Risk taking | Logistic | 325,592 | 1.02 (1.01 to 1.03) | $1.68 \times 10^{-06}$ |
| Time spent watching television | Linear | 334,630 | 0.02 (0.01 to 0.02) | $1.05 \times 10^{-26}$ |
| Liking for cabbage | Ordinal | 127,891 | 1.02 (1.01 to 1.03) | $1.94 \times 10^{-05}$ |
| Weekly usage of mobile phone | Ordinal | 281,062 | 1.02 (1.01 to 1.02) | $3.88 \times 10^{-07}$ |
| Ease of skin tanning | Ordinal | 330,266 | 0.97 (0.97 to 0.98) | $1.03 \times 10^{-17}$ |
| Mother's age at time of questionnaire | Linear | 130,898 | -0.06 (-0.09 to -0.04) | $2.25 \times 10^{-06}$ |
| Back pain experienced in last month | Logistic | 336,448 | 1.01 (1 to 1.02) | 0.05 |
| Operation on the left-side of the body | Logistic | 336,988 | 0.99 (0.97 to 1.02) | 0.58 |
| ***Smoking heaviness PRS – never smokers*** | | | | |
| Lifetime number of sexual partners | Linear | 153,616 | 0.0006 (-0.004 to 0.005) | 0.80 |
| Age at first live birth | Linear | 74,130 | -0.002 (-0.03 to 0.03) | 0.89 |
| Townsend deprivation index | Linear | 183,783 | 0.02 (0.005 to 0.03) | 0.007 |
| Takes part in a religious group | Logistic | 183,569 | 1.01 (1.00 to 1.03) | 0.17 |
| Cereal intake | Linear | 183719 | -0.003 (-0.007 to 0.001) | 0.19 |
| Risk taking | Logistic | 178,202 | 1.00 (0.99 to 1.01) | 0.98 |
| Time spent watching television | Linear | 182,824 | -0.001 (-0.006 to 0.003) | 0.58 |
| Liking for cabbage | Ordinal | 74,613 | 1.01 (1.00 to 1.03) | 0.02 |
| Weekly usage of mobile phone | Ordinal | 151,060 | 1.01 (1.00 to 1.01) | 0.26 |
| Ease of skin tanning | Ordinal | 180,056 | 1.00 (0.99 to 1.01) | 0.69 |
| Mother's age at time of questionnaire | Linear | 76,827 | 0.02 (-0.01 to 0.05) | 0.25 |

| | | | | |
|---|---|---|---|---|
| Back pain experienced in last month | Logistic | 183,749 | 0.99 (0.98 to 1.01) | 0.34 |
| Operation on the left-side of the body | Logistic | 184,005 | 0.97 (0.94 to 1.01) | 0.14 |
| ***Smoking heaviness PRS – former smokers*** | | | | |
| Lifetime number of sexual partners | Linear | 277,634 | -0.002 (-0.007 to 0.004) | 0.59 |
| Age at first live birth | Linear | 124,051 | -0.0008 (-0.05 to 0.05) | 0.97 |
| Townsend deprivation index | Linear | 336,591 | -0.002 (-0.02 to 0.01) | 0.79 |
| Takes part in a religious group | Logistic | 336,161 | 1.00 (0.98 to 1.03) | 0.81 |
| Cereal intake | Linear | 336,304 | -0.004 (-0.009 to 0.002) | 0.20 |
| Risk taking | Logistic | 325,592 | 1.00 (0.98 to 1.01) | 0.61 |
| Time spent watching television | Linear | 334,630 | 0.002 (-0.004 to 0.007) | 0.55 |
| Liking for cabbage | Ordinal | 127,891 | 1.00 (0.98 to 1.01) | 0.56 |
| Weekly usage of mobile phone | Ordinal | 281,062 | 1.00 (0.99 to 1.01) | 0.74 |
| Ease of skin tanning | Ordinal | 330,266 | 0.99 (0.98 to 1.00) | 0.21 |
| Mother’s age at time of questionnaire | Linear | 130,898 | -0.03 (-0.07 to 0.02) | 0.26 |
| Back pain experienced in last month | Logistic | 336,448 | 0.99 (0.98 to 1.02) | 0.95 |
| Operation on the left-side of the body | Logistic | 336,988 | 1.01 (0.96 to 1.06) | 0.65 |
| ***Smoking heaviness PRS – current smokers*** | | | | |
| Lifetime number of sexual partners | Linear | 277,634 | -0.002 (-0.01 to 0.009) | 0.69 |
| Age at first live birth | Linear | 124,051 | -0.09 (-0.19 to -0.003) | 0.04 |
| Townsend deprivation index | Linear | 336,591 | -0.01 (-0.05 to 0.03) | 0.55 |
| Takes part in a religious group | Logistic | 336,161 | 1.02 (0.96 to 1.08) | 0.58 |
| Cereal intake | Linear | 336,304 | -0.004 (-0.01 to 0.007) | 0.50 |
| Risk taking | Logistic | 325,592 | 1.00 (0.98 to 1.02) | 0.98 |
| Time spent watching television | Linear | 334,630 | -0.002 (-0.01 to 0.01) | 0.77 |
| Liking for cabbage | Ordinal | 127,891 | 0.98 (0.94 to 1.02) | 0.31 |
| Weekly usage of mobile phone | Ordinal | 281,062 | 1.00 (0.98 to 1.02) | 0.77 |
| Ease of skin tanning | Ordinal | 330,266 | 0.99 (0.97 to 1.01) | 0.21 |
| Mother’s age at time of questionnaire | Linear | 130,898 | 0.04 (-0.05 to 0.12) | 0.40 |
| Back pain experienced in last month | Logistic | 336,448 | 0.98 (0.95 to 1.01) | 0.24 |

| | | | | |
|---|---|---|---|---|
| Operation on the left-side of the body | Logistic | 336,988 | 0.95 (0.87 to 1.05) | 0.32 |
| ***Smoking initiation PRS*** | | | | |
| Lifetime number of sexual partners | Linear | 277,634 | 0.02 (0.02 to 0.02) | $6.39x10^{-29}$ |
| Age at first live birth | Linear | 124,051 | -0.08 (-0.11 to -0.06) | $8.26x10^{-11}$ |
| Townsend deprivation index | Linear | 336,591 | 0.04 (0.03 to 0.05) | $7.15x10^{-16}$ |
| Takes part in a religious group | Logistic | 336,161 | 0.97 (0.96 to 0.98) | $1.29x10^{-07}$ |
| Cereal intake | Linear | 336,304 | -0.01 (-0.01 to -0.01) | $3.46x10^{-08}$ |
| Risk taking | Logistic | 325,592 | 1.03 (1.02 to 1.03) | $2.93x10^{-10}$ |
| Time spent watching television | Linear | 334,630 | 0.01 (0.01 to 0.02) | $6.18x10^{-14}$ |
| Liking for cabbage | Ordinal | 127,891 | 1.02 (1.01 to 1.03) | $2.49x10^{-06}$ |
| Weekly usage of mobile phone | Ordinal | 281,062 | 1.02 (1.01 to 1.03) | $3.25x10^{-08}$ |
| Ease of skin tanning | Ordinal | 330,266 | 0.99 (0.98 to 0.99) | $6.34x10^{-06}$ |
| Mother's age at time of questionnaire | Linear | 130,898 | -0.07 (-0.1 to -0.05) | $1.72x10^{-08}$ |
| Back pain experienced in last month | Logistic | 336,448 | 1.01 (1 to 1.02) | 0.19 |
| Operation on the left-side of the body | Logistic | 336,988 | 0.96 (0.94 to 0.99) | 0.005 |

For linear regression models the effect estimate is beta and for logistic and ordinal regressions this is the odds ratio. OR=odds ratio, CI=confidence interval, PRS=polygenic risk score.

**Supplementary Table S9. Summary statistics for ALSPAC sample with data available**

| Phenotype | Sample | N | Mean (SD) or percentage (%) |
|---|---|---|---|
| Ever smoked | Mother | 12,997 | 50.90% |
| | Father/ Partner | 9,820 | 55.20% |
| Smoking heaviness (CPD, for ever smokers) | Mother (T1) | 11,568 | None=77.32%<br>1 to 4=3.99%<br>5 to 9=4.47%<br>10 to 14=5.81%<br>15 to 19=4.09%<br>20 to 24=3.09%<br>25 to 29=0.73%<br>30+=0.52% |
| | Mother (T2) | 11,018 | None=75.75%<br>1 to 4=3.82%<br>5 to 9=4.47%<br>10 to 14=5.81%<br>15 to 19=4.92%<br>20 to 24=3.96%<br>25 to 29=0.93%<br>30+=0.35% |
| | Father/ Partner | 7,001 | None=72.58%<br>1 to 4=3.34%<br>5 to 9=3.57%<br>10 to 14=5.78%<br>15 to 19=5.70%<br>20 to 24=6.01%<br>25 to 29=1.81%<br>30+=1.20% |
| Age at first time point (years) | Mother | 10,135 | 27.75 (4.92) |
| | Father/ Partner | 8,121 | 30.64 (5.70) |
| | Child | 11,878 | 0.10 (0.06) |
| Sex (females) | Child | 13,923 | 48.38% |
| BMI | Mother | 4,557 | 26.57 (5.13) |
| | Father/ Partner | 1,884 | 27.48 (3.89) |
| | Child | 7,567 | 16.16 (1.88) |
| Body fat percentage | Mother | 4,412 | 36.75 (8.08) |

| | | | |
|---|---|---|---|
| | Father/ Partner | 1,758 | 26.61 (6.82) |
| | Child | 6,843 | 23.14 (9.06) |
| Wheeze | Mother | 12,223 | 16.89% |
| | Father/ Partner | 8,443 | 18.44% |
| | Child (T1) | 11,314 | 21.49% |
| | Child (T2) | 10,157 | 19.32% |
| C-reactive protein | Mother | 3,889 | 1.52 (1.55) |
| | Father/ Partner | 1,713 | 1.53 (1.32) |
| | Child | 4,392 | 0.33 (0.35) |
| Overall health | Mother (T1) | 11,436 | Hardly ever well=0.70%<br>Often unwell=4.74%<br>Mostly well=63.21%<br>Always well=31.35% |
| | Mother (T2) | 11,116 | Hardly ever well=0.76%<br>Often unwell=4.17%<br>Mostly well=62.71%<br>Always well=32.35% |
| | Father/ Partner (T1) | 8,285 | Rarely well=0.22%<br>Often unwell=1.88%<br>Mostly healthy=44.39%<br>Always healthy=53.51% |
| | Father/ Partner (T2) | 7,059 | Hardly ever well=0.45%<br>Often feel unwell=2.42%<br>Mostly feel well=52.61%<br>Always fit and well=44.51% |
| | Child (T1) | 12,094 | Almost always unwell=0.12%<br>Sometimes quite ill=0.21%<br>Healthy=19.36%<br>Very healthy=80.31% |
| | Child (T2) | 11,252 | Mostly unwell=0.93%<br>Sometimes quite ill=2.58%<br>Minor problems=36.61%<br>Very healthy=59.88% |
| Gamma glutamyl transferase | Child | 4,703 | 16.80 (5.12) |
| Seen GP for nerves/anxiety | Mother (T1) | 11,088 | No=79.25%<br>Yes, did not see GP=16.41%<br>Yes, saw GP=4.35% |
| | Mother (T2) | 10,018 | No=81.30%<br>Yes, did not see GP=13.73% |

| | | | |
|---|---|---|---|
| | | | Yes, saw GP=4.97% |
| | Father/<br>Partner (T1) | 7,046 | No=86.19%<br>Yes, did not see GP=12.18%<br>Yes, saw GP=1.63% |
| | Father/<br>Partner (T2) | 6,123 | No=85.20%<br>Yes, did not see GP=13.10%<br>Yes, saw GP=1.70% |
| | Child | 8,002 | 33.87% |
| Seen GP for depression | Mother (T1) | 11,088 | No=68.70%<br>Yes, did not see GP=23.34%<br>Yes, saw GP=7.95% |
| | Mother (T2) | 10,063 | No=75.81%<br>Yes, did not see GP=16.28%<br>Yes, saw GP=7.91% |
| | Father/<br>Partner (T1) | 7,044 | No=89.34%<br>Yes, did not see GP=9.58%<br>Yes, saw GP=1.08% |
| | Father/<br>Partner (T2) | 6,123 | No=88.63%<br>Yes, did not see GP=9.83%<br>Yes, saw GP=1.54% |
| Number of treatments/ medications taken | Mother (T1) | 11,612 | 1.29 (1.58) |
| | Mother (T2) | 7,662 | 1.94 (1.11) |
| | Father/<br>Partner (T1) | 4,325 | 1.30 (1.03) |
| | Father/<br>Partner (T2) | 3,027 | 1.61 (1.33) |
| | Child (T1) | 12,159 | 1.72 (1.54) |
| | Child (T2) | 10,232 | 4.03 (1.53) |
| Alcohol drunk on typical drinking day | Mother | 12,982 | Never=8.44%<br><1 per week=37.60%<br>>1 per week=42.78%<br>1 to 2 per day=9.48%<br>3 to 9 per day=1.58%<br>>10 per day=0.12% |
| | Father/<br>Partner | 9,833 | Never=3.77%<br><1 per week=21.42%<br>>1 per week=51.66%<br>1 to 2 per day=16.98%<br>3 to 9 per day=5.75%<br>>10 per day=0.42% |
| Age at first pregnancy | Mother | 13,093 | 24.33 (4.99) |

| | | | |
|---|---|---|---|
| Townsend deprivation index | Mother (T1) | 7,991 | 1=25.83%<br>2=14.52%<br>3=18.71%<br>4=26.63%<br>5=14.32% |
| | Mother (T2) | 10,583 | 1=27.34%<br>2=14.76%<br>3=19.48%<br>4=25.22%<br>5=13.20% |
| Attends a place of worship | Mother | 11,974 | Not at all=56.56%<br>>1 per year=29.19%<br>>1 per month=6.91%<br>>1 per week=7.34% |
| | Father/ Partner | 9,543 | Not at all=63.38%<br>>1 per year=26.23%<br>>1 per month=4.30%<br>>1 per week=6.10% |
| | Child | 7,327 | No=54.24%<br>Yes, sometimes=29.68%<br>Yes, often=16.08% |
| Oat cereal intake | Mother (T1) | 11,988 | Never/rarely=43.71%<br>Once in 2 weeks=15.27%<br>1 to 3 times per week=17.16%<br>4 to 7 times per week=21.76%<br>>1 per day=2.10% |
| | Mother (T2) | 9,433 | Never/rarely=61.44%<br>Once in 2 weeks=16.07%<br>1 to 3 times per week=12.70%<br>4 to 7 times per week=9.39%<br>>1 per day=0.39% |
| | Father/ Partner | 5,031 | Never/rarely=62.41%<br>Once in 2 weeks=15.11%<br>1 to 3 times per week=11.51%<br>4 to 7 times per week=10.55%<br>>1 per day=0.42% |
| | Child | 8,984 | Never/rarely=53.52%<br>Once in 2 weeks=17.52%<br>1 to 3 times per week=21.26%<br>4 to 7 times per week=7.50%<br>>1 per day=0.20% |
| Wholegrain/bran cereal intake | Mother (T1) | 11,988 | Never/rarely=32.56%<br>Once in 2 weeks=13.96%<br>1 to 3 times per week=24.02%<br>4 to 7 times per week=27.47%<br>>1 per day=1.99% |

| | | | |
|---|---|---|---|
| | Mother (T2) | 9,443 | Never/rarely=34.80%<br>Once in 2 weeks=17.28%<br>1 to 3 times per week=21.91%<br>4 to 7 times per week=25.08%<br>>1 per day=0.92% |
| | Father/ Partner | 5,030 | Never/rarely=42.03%<br>Once in 2 weeks=17.26%<br>1 to 3 times per week=20.28%<br>4 to 7 times per week=19.58%<br>>1 per day=0.85% |
| | Child | 9,176 | Never/rarely=21.71%<br>Once in 2 weeks=13.35%<br>1 to 3 times per week=39.28%<br>4 to 7 times per week=24.67%<br>>1 per day=0.99% |
| Other cereal intake | Mother (T1) | 11,988 | Never/rarely=37.99%<br>Once in 2 weeks=19.53%<br>1 to 3 times per week=26.40%<br>4 to 7 times per week=15.28%<br>>1 per day=0.81% |
| | Mother (T2) | 9,443 | Never/rarely=38.95%<br>Once in 2 weeks=18.88%<br>1 to 3 times per week=25.44%<br>4 to 7 times per week=15.94%<br>>1 per day=0.80% |
| | Father/ Partner | 5,031 | Never/rarely=34.47%<br>Once in 2 weeks=20.57%<br>1 to 3 times per week=26.22%<br>4 to 7 times per week=17.25%<br>>1 per day=1.13% |
| | Child | 9,386 | Never/rarely=7.56%<br>Once in 2 weeks=7.89<br>1 to 3 times per week=42.82%<br>4 to 7 times per week=40.02%<br>>1 per day=1.70% |
| Risk taking (higher = avoids risks more) | Child (T1) | 10,021 | Never=18.71%<br>Hardly ever=31.27%<br>Sometimes=37.14%<br>Often=10.09%<br>Very often=2.78% |
| | Child (T2) | 9,891 | Never=15.44%<br>Hardly ever=29.08%<br>Sometimes=40.34%<br>Often=12.01%<br>Very often=3.13% |
| Time spent watching television (weekday) | Mother | 6,981 | 2.38 (1.37) |

| | | | |
|---|---|---|---|
| | Father/ Partner | 3,637 | 2.30 (1.32) |
| | Child | 9,913 | Not at all=2.10%<br><1 hour=26.44%<br>1 to 2 hours=46.08%<br>>2 hours=25.38% |
| Time spent watching television (weekend day) | Mother | 6,491 | 3.15 (1.75) |
| | Father/ Partner | 3,427 | 3.31 (1.82) |
| | Child | 9,830 | Not at all=3.34%<br><1 hour=25.34%<br>1 to 2 hours=43.62%<br>>2 hours=27.70% |
| Cabbage intake | Mother (T1) | 11,988 | Never/rarely=9.92%<br>Once in 2 weeks=19.49%<br>1 to 3 times per week=60.07%<br>4 to 7 times per week=10.23%<br>>1 per day=0.30% |
| | Mother (T2) | 9,428 | Never/rarely=7.24%<br>Once in 2 weeks=17.02%<br>1 to 3 times per week=62.47%<br>4 to 7 times per week=13.06%<br>>1 per day=0.20% |
| | Father/ Partner | 9,798 | Never/rarely=12.05%<br>Once in 2 weeks=17.03%<br>1 to 3 times per week=53.50%<br>4 to 7 times per week=16.72%<br>>1 per day=0.69% |
| | Child (T1) | 9,891 | Never/rarely=31.57%<br>Once in 2 weeks=16.78%<br>1 to 3 times per week=43.88%<br>4 to 7 times per week=7.56%<br>>1 per day=0.20% |
| | Child (T2) | 9,501 | Never/rarely=26.68%<br>Once in 2 weeks=14.20%<br>1 to 3 times per week=49.84%<br>4 to 7 times per week=9.08%<br>>1 per day=0.20% |
| Mobile phone usage | Mother | 3,635 | <1 per week=40.69%<br>1 to 3 times per week=26.19%<br>4 to 6 times per week=11.77%<br>>1 per day=21.35% |
| | Father/ Partner | 2,243 | <1 per week=17.12%<br>1 to 3 times per week=18.86%<br>4 to 6 times per week=15.07%<br>>1 per day=48.95% |

| | | | |
|---|---|---|---|
| | Child (weekday) | 6,099 | Not at all=65.55%<br><15 minutes=30.07%<br>15 to 30 minutes=3.16%<br>>30 minutes=1.21% |
| | Child (weekend day) | 6,440 | Not at all=27.03%<br><15 minutes=58.73%<br>15 to 30 minutes=9.92%<br>>30 minutes=4.32% |
| Mother's age at time of questionnaire | Mother | 10,135 | 54.75 (8.28) |
| | Father/ Partner | 7,209 | 57.15 (8.58) |
| Back pain | Mother (T1) | 11,555 | 67.65% |
| | Mother (T2) | 11,088 | 59.60% |
| | Father/ Partner | 8,451 | 46.92% |

CPD=cigarettes per day. T1 = time point 1, T2 = time point 2

**Supplementary Table S10. Associations between the polygenic risk scores for lifetime smoking score, smoking heaviness and smoking initiation and plausible outcomes in ALSPAC**

| Outcome | Model | Sample | N | Beta or OR (95% CI) | P-value |
|---|---|---|---|---|---|
| ***Lifetime smoking score PRS*** | | | | | |
| BMI | Linear | Mother | 3,176 | 0.22 (0.04 to 0.39) | 0.02 |
| | | Father/ Partner | 1,100 | 0.23 (0.007 to 0.46) | 0.04 |
| | | Child | 5,474 | 0.05 (-0.002 to 0.10) | 0.06 |
| Body fat percentage | Linear | Mother | 3,097 | 0.20 (-0.09 to 0.49) | 0.18 |
| | | Father/ Partner | 1,018 | 0.23 (-0.18 to 0.64) | 0.26 |
| | | Child | 5,078 | 0.25 (0.02 to 0.48) | 0.03 |
| Wheeze | Logistic | Mother | 6,720 | 1.07 (1.01 to 1.15) | 0.03 |
| | | Father/ Partner | 1,386 | 0.95 (0.82 to 1.10) | 0.49 |
| | | Child (T1) | 6,716 | 0.99 (0.94 to 1.05) | 0.84 |
| | | Child (T2) | 6,081 | 1.06 (0.99 to 1.13) | 0.09 |
| C-reactive protein | Linear | Mother | 2,773 | -0.008 (-0.05 to 0.03) | 0.67 |
| | | Father/ Partner | 1,015 | 0.03 (-0.03 to 0.09) | 0.37 |
| | | Child | 3,570 | 0.006 (-0.03 to 0.04) | 0.69 |
| Overall health | Ordinal | Mother (T1) | 6,675 | 0.95 (0.90 to 1.00) | 0.05 |
| | | Mother (T2) | 3,029 | 0.98 (0.93 to 1.03) | 0.43 |
| | | Father/ Partner (T1) | 1,307 | 0.91 (0.82 to 1.02) | 0.10 |
| | | Father/ Partner (T2) | 1,279 | 0.89 (0.80 to 1.00) | 0.05 |
| | | Child (T1) | 6,864 | 1.01 (0.95 to 1.07) | 0.70 |
| | | Child (T2) | 6,689 | 1.02 (0.97 to 1.07) | 0.42 |
| Gamma glutamyl transferase | Linear | Child | 3,811 | 0.08 (-0.08 to 0.24) | 0.35 |
| Seen GP for nerves/anxiety | Ordinal | Mother (T1) | 6,452 | 1.00 (0.94 to 1.06) | 0.93 |
| | | Mother (T2) | 5,593 | 1.04 (0.97 to 1.12) | 0.22 |
| | | Father/ | 1,277 | 1.15 (0.97 to 1.36) | 0.10 |

| | | | | | |
|---|---|---|---|---|---|
| | | Partner (T1) | | | |
| | | Father/ Partner (T2) | 1,203 | 1.13 (0.96 to 1.33) | 0.15 |
| | Logistic | Child | 5,381 | 0.94 (0.89 to 1.00) | 0.04 |
| Seen GP for depression | Ordinal | Mother (T1) | 6,452 | 1.05 (1.00 to 1.11) | 0.07 |
| | | Mother (T2) | 5,617 | 1.09 (1.02 to 1.16) | 0.01 |
| | | Father/ Partner (T1) | 1,277 | 1.09 (0.90 to 1.34) | 0.37 |
| | | Father (T2) | 1,203 | 1.02 (0.85 to 1.23) | 0.81 |
| Number of treatments/ medications taken | Linear | Mother (T1) | 6,662 | -0.02 (-0.04 to 0.008) | 0.19 |
| | | Mother (T2) | 4,343 | -0.004 (-0.03 to 0.03) | 0.81 |
| | | Father/ Partner (T1) | 901 | -0.03 (-0.10 to 0.04) | 0.37 |
| | | Father/ Partner (T2) | 831 | 0.008 (-0.06 to 0.07) | 0.82 |
| | | Child (T1) | 6,893 | -0.009 (-0.03 to 0.01) | 0.46 |
| | | Child (T2) | 6,034 | 0.03 (-0.009 to 0.07) | 0.13 |
| Alcohol drunk on typical drinking day | Ordinal | Mother | 7,167 | 1.02 (0.98 to 1.07) | 0.30 |
| | | Father/ Partner | 1,467 | 1.08 (0.98 to 1.19) | 0.13 |
| ***Smoking heaviness PRS – never smokers*** | | | | | |
| BMI | Linear | Mother | 1,885 | -0.13 (-0.35 to 0.10) | 0.26 |
| | | Father/ Partner | 594 | 0.18 (-0.12 to 0.49) | 0.23 |
| | | Child | 5,474 | 0.03 (-0.02 to 0.08) | 0.18 |
| Body fat percentage | Linear | Mother | 1,842 | -0.27 (-0.64 to 0.10) | 0.15 |
| | | Father/ Partner | 554 | 0.22 (-0.34 to 0.77) | 0.44 |
| | | Child | 5,078 | 0.13 (-0.10 to 0.36) | 0.28 |
| Wheeze | Logistic | Mother | 3,460 | 0.97 (0.88 to 1.07) | 0.54 |
| | | Father/ Partner | 769 | 0.97 (0.79 to 1.19) | 0.74 |

| | | | | | |
|---|---|---|---|---|---|
| | | Child (T1) | 6,716 | 0.98 (0.93 to 1.05) | 0.62 |
| | | Child (T2) | 6,081 | 1.03 (0.97 to 1.10) | 0.33 |
| C-reactive protein | Linear | Mother | 1,662 | 0.007 (-0.04 to 0.05) | 0.77 |
| | | Father/ Partner | 544 | 0.01 (-0.07 to 0.10) | 0.75 |
| | | Child | 3,570 | 0.03 (-0.001 to 0.06) | 0.06 |
| Overall health | Ordinal | Mother (T1) | 3,392 | 1.01 (0.94 to 1.08) | 0.84 |
| | | Mother (T2) | 3,365 | 1.03 (0.96 to 1.10) | 0.46 |
| | | Father/ Partner (T1) | 710 | 0.91 (0.78 to 1.06) | 0.23 |
| | | Father/ Partner (T2) | 699 | 1.00 (0.86 to 1.17) | 0.97 |
| | | Child (T1) | 6,864 | 1.01 (0.95 to 1.08) | 0.71 |
| | | Child (T2) | 6,689 | 0.98 (0.93 to 1.03) | 0.36 |
| Gamma glutamyl transferase | Linear | Child | 3,811 | 0.13 (-0.03 to 0.29) | 0.12 |
| Seen GP for nerves/anxiety | Ordinal | Mother (T1) | 3,358 | 0.98 (0.89 to 1.07) | 0.60 |
| | | Mother (T2) | 2,996 | 1.03 (0.93 to 1.13) | 0.62 |
| | | Father/ Partner (T1) | 696 | 1.15 (0.90 to 1.46) | 0.27 |
| | | Father/ Partner (T2) | 667 | 1.00 (0.80 to 1.25) | 0.99 |
| | Logistic | Child | 5,381 | 1.01 (0.96 to 1.07) | 0.70 |
| Seen GP for depression | Ordinal | Mother (T1) | 3,358 | 1.00 (0.92 to 1.08) | 0.97 |
| | | Mother (T2) | 3,011 | 1.01 (0.93 to 1.11) | 0.77 |
| | | Father/ Partner (T1) | 696 | 1.12 (0.81 to 1.57) | 0.49 |
| | | Father/ Partner (T2) | 667 | 0.94 (0.71 to 1.25) | 0.66 |
| Number of treatments/ medications taken | Linear | Mother (T1) | 3,436 | 0.02 (-0.01 to 0.05) | 0.26 |

| | | | | | |
|---|---|---|---|---|---|
| | | Mother (T2) | 2,295 | -0.05 (-0.09 to -0.008) | 0.02 |
| | | Father/ Partner (T1) | 499 | -0.04 (-0.12 to 0.05) | 0.39 |
| | | Father/ Partner (T2) | 450 | -0.0004 (-0.10 to 0.09) | 0.99 |
| | | Child (T1) | 6,893 | -0.005 (-0.03 to 0.02) | 0.70 |
| | | Child (T2) | 6,034 | -0.009 (-0.05 to 0.03) | 0.63 |
| Alcohol drunk on typical drinking day | Ordinal | Mother | 3,682 | 0.99 (0.93 to 1.05) | 0.81 |
| | | Father/ Partner | 819 | 1.00 (0.88 to 1.15) | 0.96 |
| ***Smoking heaviness PRS – ever smokers*** | | | | | |
| BMI | Linear | Mother | 1,237 | -0.10 (-0.38 to 0.19) | 0.52 |
| | | Father/ Partner | 425 | -0.02 (-0.40 to 0.35) | 0.91 |
| Body fat percentage | Linear | Mother | 1,202 | -0.14 (-0.61 to 0.34) | 0.57 |
| | | Father/ Partner | 390 | -0.07 (-0.73 to 0.60) | 0.85 |
| Wheeze | Logistic | Mother | 3,133 | 1.09 (1.00 to 1.19) | 0.06 |
| | | Father/ Partner | 575 | 0.87 (0.70 to 1.07) | 0.19 |
| C-reactive protein | Linear | Mother | 1,061 | 0.07 (0.01 to 0.14) | 0.02 |
| | | Father/ Partner | 395 | -0.04 (-0.14 to 0.06) | 0.41 |
| Overall health | Ordinal | Mother (T1) | 3,036 | 0.96 (0.89 to 1.03) | 0.27 |
| | | Mother (T2) | 2,948 | 0.98 (0.91 to 1.06) | 0.63 |
| | | Father/ Partner (T1) | 553 | 0.88 (0.74 to 1.05) | 0.15 |
| | | Father/ Partner (T2) | 536 | 1.10 (0.92 to 1.33) | 0.31 |
| Seen GP for nerves/anxiety | Ordinal | Mother (F) | 2,945 | 1.02 (0.94 to 1.11) | 0.67 |
| | | Mother (G) | 2,511 | 0.99 (0.90 to 1.09) | 0.87 |
| | | Father/ Partner (T1) | 537 | 0.81 (0.62 to 1.07) | 0.13 |

| | | | | | |
|---|---|---|---|---|---|
| | | Father/ Partner (T2) | 493 | 0.87 (0.67 to 1.13) | 0.29 |
| Seen GP for depression | Ordinal | Mother (T1) | 2,945 | 1.02 (0.95 to 1.10) | 0.60 |
| | | Mother (T2) | 2,517 | 0.97 (0.88 to 1.06) | 0.44 |
| | | Father/ Partner (T1) | 537 | 0.94 (0.69 to 1.28) | 0.70 |
| | | Father/ Partner (T2) | 493 | 1.01 (0.78 to 1.31) | 0.92 |
| Number of treatments/ medications taken | Linear | Mother (T1) | 3,076 | -0.01 (-0.05 to 0.02) | 0.43 |
| | | Mother (T2) | 1,982 | -0.02 (-0.06 to 0.02) | 0.43 |
| | | Father/ Partner (T1) | 375 | -0.14 (-0.24 to -0.03) | 0.01 |
| | | Father/ Partner (T2) | 350 | -0.03 (-0.12 to 0.07) | 0.59 |
| Alcohol drunk on typical drinking day | Ordinal | Mother | 3,454 | 0.92 (0.86 to 0.98) | 0.006 |
| | | Father/ Partner | 642 | 0.99 (0.85 to 1.16) | 0.94 |
| ***Smoking initiation PRS*** | | | | | |
| BMI | Linear | Mother | 3,176 | 0.36 (0.19 to 0.54) | 0.00005 |
| | | Father/ Partner | 1,100 | 0.22 (-0.006 to 0.44) | 0.06 |
| | | Child | 5,474 | 0.04 (-0.009 to 0.09) | 0.11 |
| Body fat percentage | Linear | Mother | 3,097 | 0.34 (0.05 to 0.62) | 0.02 |
| | | Father/ Partner | 1,018 | 0.16 (-0.25 to 0.56) | 0.44 |
| | | Child | 5,078 | 0.12 (-0.11 to 0.35) | 0.31 |
| Wheeze | Logistic | Mother | 6,720 | 1.02 (0.96 to 1.09) | 0.48 |
| | | Father/ Partner | 1,386 | 1.03 (0.89 to 1.19) | 0.69 |
| | | Child (T1) | 6,716 | 1.00 (0.94 to 1.06) | 0.97 |
| | | Child (T2) | 6,081 | 1.06 (1.00 to 1.13) | 0.07 |
| C-reactive protein | Linear | Mother | 2,773 | -0.01 (-0.05 to 0.02) | 0.50 |
| | | Father/ Partner | 1,015 | 0.03 (-0.04 to 0.09) | 0.42 |

| | | | | | |
|---|---|---|---|---|---|
| | | Child | 3,570 | 0.02 (-0.01 to 0.05) | 0.27 |
| Overall health | Ordinal | Mother (T1) | 6,575 | 0.97 (0.92 to 1.02) | 0.23 |
| | | Mother (T2) | 6,462 | 1.03 (0.98 to 1.08) | 0.31 |
| | | Father/ Partner (T1) | 1,307 | 0.94 (0.84 to 1.05) | 0.26 |
| | | Father/ Partner (T2) | 1,279 | 0.98 (0.88 to 1.09) | 0.73 |
| | | Child (T1) | 6,864 | 1.03 (0.97 to 1.09) | 0.34 |
| | | Child (T2) | 6,689 | 1.02 (0.97 to 1.07) | 0.43 |
| Gamma glutamyl transferase | Linear | Child | 3,811 | 0.10 (-0.06 to 0.26) | 0.22 |
| Seen GP for nerves/anxiety | Ordinal | Mother (T1) | 6,452 | 1.03 (0.96 to 1.09) | 0.42 |
| | | Mother (T2) | 5,593 | 1.01 (0.94 to 1.08) | 0.75 |
| | | Father/ Partner (T2) | 1,277 | 1.00 (0.84 to 1.18) | 0.98 |
| | | Father/ Partner (T2) | 1,203 | 0.99 (0.84 to 1.16) | 0.87 |
| | Logistic | Child | 5,381 | 0.92 (0.87 to 0.97) | 0.004 |
| Seen GP for depression | Ordinal | Mother (T1) | 6,452 | 1.04 (0.98 to 1.10) | 0.16 |
| | | Mother (T2) | 5,617 | 1.02 (0.96 to 1.09) | 0.51 |
| | | Father/ Partner (T1) | 1,277 | 0.90 (0.73 to 1.11) | 0.32 |
| | | Father/ Partner (T2) | 1,203 | 1.08 (0.90 to 1.31) | 0.41 |
| Number of treatments/ medications taken | Linear | Mother (T1) | 6,662 | -0.007 (-0.03 to 0.02) | 0.55 |
| | | Mother (T2) | 4,343 | -0.003 (-0.03 to 0.03) | 0.85 |
| | | Father/ Partner (T1) | 901 | 0.003 (-0.06 to 0.07) | 0.93 |
| | | Father (T2) | 831 | -0.02 (-0.08 to 0.05) | 0.57 |

| | | | | | |
|---|---|---|---|---|---|
| | | Child (T1) | 6,893 | -0.01 (-0.03 to 0.01) | 0.42 |
| | | Child (T2) | 6,034 | 0.01 (-0.03 to 0.05) | 0.56 |
| Alcohol drunk on typical drinking day | Ordinal | Mother | 7,167 | 1.06 (1.02 to 1.11) | 0.006 |
| | | Father | 1,467 | 1.09 (0.99 to 1.21) | 0.08 |

For linear regression models the effect estimate is beta and for logistic and ordinal regressions this is the odds ratio. OR=odds ratio, CI=confidence interval, PRS=polygenic risk score, BMI=body mass index, COPD=chronic obstructive pulmonary disease, GP=general practitioner. T1 = time point 1, T2 = time point 2

**Supplementary Table S11. Associations between the polygenic risk scores for lifetime smoking score, smoking heaviness and smoking initiation and implausible outcomes in ALSPAC**

| Outcome | Model | Sample | N | Beta or OR (95% CI) | P-value |
|---|---|---|---|---|---|
| ***Lifetime smoking score PRS*** | | | | | |
| Age at first pregnancy | Linear | Mother | 7,199 | -0.19 (-0.28 to -0.10) | $5.85 \times 10^{-05}$ |
| Townsend deprivation index | Ordinal | Mother (T1) | 4,376 | 1.06 (1.01 to 1.12) | 0.02 |
| | | Mother (T2) | 6,174 | 1.05 (1.00 to 1.09) | 0.05 |
| Attends a place of worship | Ordinal | Mother | 2,990 | 0.91 (0.87 to 0.96) | 0.0002 |
| | | Father | 1,442 | 0.98 (0.80 to 1.12) | 0.77 |
| | | Child | 5,110 | 0.90 (0.85 to 0.95) | 0.00008 |
| Oat cereal intake | Ordinal | Mother (T1) | 6,712 | 0.96 (0.92 to 1.01) | 0.09 |
| | | Mother (T2) | 5,662 | 0.91 (0.85 to 0.98) | 0.02 |
| | | Father | 1,155 | 0.90 (0.80 to 1.01) | 0.06 |
| | | Child | 5,671 | 0.97 (0.92 to 1.01) | 0.17 |
| Wholegrain/bran cereal intake | Ordinal | Mother (T1) | 6,712 | 0.92 (0.86 to 0.96) | 0.0003 |
| | | Mother (T2) | 5,662 | 0.94 (0.89 to 0.98) | 0.007 |
| | | Father | 1,156 | 0.98 (0.88 to 1.08) | 0.65 |
| | | Child | 5,776 | 0.97 (0.92 to 1.01) | 0.14 |
| Other cereal intake | Ordinal | Mother (T1) | 6,712 | 1.05 (1.00 to 1.09) | 0.04 |
| | | Mother (T2) | 5,662 | 0.98 (0.94 to 1.03) | 0.50 |
| | | Father | 1,149 | 0.95 (0.86 to 1.06) | 0.36 |
| | | Child | 5,901 | 1.01 (0.96 to 1.06) | 0.76 |
| Risk taking (higher = avoids risks more) | Ordinal | Child (T1) | 6,023 | 0.92 (0.88 to 0.96) | 0.0005 |
| | | Child (T2) | 5,991 | 0.96 (0.92 to 1.01) | 0.08 |
| Time spent watching television (weekday) | Linear | Mother | 4,426 | 0.05 (0.02 to 0.08) | 0.0006 |
| | | Father | 1,066 | 0.03 (-0.03 to 0.09) | 0.35 |
| | Ordinal | Child | 6,072 | 1.03 (0.99 to 1.08) | 0.17 |
| Time spent watching television (weekend day) | Linear | Mother | 4,129 | 0.03 (0.006 to 0.06) | 0.02 |
| | | Father | 1,019 | 0.04 (-0.02 to 0.10) | 0.19 |
| | Ordinal | Child | 6,033 | 1.06 (1.01 to 1.11) | 0.01 |
| Cabbage intake | Ordinal | Mother (T1) | 6,712 | 1.01 (0.97 to 1.06) | 0.55 |

| | | | | | |
|---|---|---|---|---|---|
| | | Mother (T2) | 5,661 | 1.03 (0.97 to 1.08) | 0.35 |
| | | Father | 1,463 | 1.04 (0.95 to 1.15) | 0.40 |
| | | Child (T1) | 6,058 | 1.06 (1.01 to 1.11) | 0.02 |
| | | Child (T2) | 5,973 | 1.007 (0.96 to 1.06) | 0.78 |
| Mobile phone usage | Ordinal | Mother | 2,306 | 1.08 (1.00 to 1.16) | 0.06 |
| | | Father | 630 | 0.98 (0.84 to 1.14) | 0.75 |
| | | Child (weekday) | 4,208 | 1.06 (1.00 to 1.13) | 0.06 |
| | | Child (weekend day) | 4,444 | 1.05 (0.99 to 1.11) | 0.09 |
| Mother's age at time of questionnaire | Linear | Mother | 5,688 | -0.17 (-0.32 to -0.03) | 0.02 |
| | | Father | 1,217 | -0.31 (-0.62 to -0.002) | 0.05 |
| Back pain | Logistic | Mother (T1) | 6,643 | 1.03 (0.98 to 1.08) | 0.26 |
| | | Mother (T2) | 6,652 | 1.00 (0.94 to 1.04) | 0.69 |
| | | Father | 1,388 | 0.98 (0.88 to 1.09) | 0.66 |
| ***Smoking heaviness PRS – never smokers*** | | | | | |
| Age at first pregnancy | Linear | Mother | 3,690 | 0.03 (-0.08 to 0.15) | 0.56 |
| Townsend deprivation index | Ordinal | Mother (T1) | 2,176 | 1.03 (0.95 to 1.11) | 0.48 |
| | | Mother (T2) | 3,173 | 1.05 (0.99 to 1.12) | 0.12 |
| Attends a place of worship | Ordinal | Mother | 3,426 | 1.01 (0.95 to 1.07) | 0.78 |
| | | Father | 808 | 1.05 (0.92 to 1.20) | 0.49 |
| | | Child | 5,110 | 0.99 (0.94 to 1.05) | 0.79 |
| Oat cereal intake | Ordinal | Mother (T1) | 3,451 | 0.98 (0.92 to 1.04) | 0.51 |
| | | Mother (T2) | 3,008 | 0.98 (0.91 to 1.05) | 0.52 |
| | | Father | 637 | 1.07 (0.92 to 1.24) | 0.40 |
| | | Child | 5,671 | 1.009 (0.96 to 1.06) | 0.72 |
| Wholegrain/bran cereal intake | Ordinal | Mother (T1) | 3,451 | 0.99 (0.93 to 1.05) | 0.67 |
| | | Mother (T2) | 3,008 | 1.00 (0.94 to 1.06) | 0.96 |
| | | Father | 638 | 1.04 (0.90 to 1.20) | 0.56 |
| | | Child | 5,776 | 0.99 (0.94 to 1.03) | 0.54 |
| Other cereal intake | Ordinal | Mother (T1) | 3,451 | 0.95 (0.90 to 1.01) | 0.11 |
| | | Mother (T2) | 3,008 | 0.97 (0.91 to 1.04) | 0.38 |
| | | Father | 635 | 0.99 (0.85 to 1.13) | 0.78 |

| | | | | | |
|---|---|---|---|---|---|
| | | Child | 5,901 | 1.0009 (0.95 to 1.05) | 0.97 |
| Risk taking (higher = avoids risks more) | Ordinal | Child (T1) | 6,023 | 0.98 (0.93 to 1.02) | 0.30 |
| | | Child (T2) | 5,991 | 1.00 (0.96 to 1.05) | 0.94 |
| Time spent watching television (weekday) | Linear | Mother | 2,397 | 0.006 (-0.03 to 0.04) | 0.76 |
| | | Father | 595 | 0.01 (-0.07 to 0.09) | 0.75 |
| | Ordinal | Child | 6,072 | 1.00 (0.95 to 1.05) | 0.95 |
| Time spent watching television (weekend day) | Linear | Mother | 2,238 | 0.02 (-0.02 to 0.06) | 0.33 |
| | | Father | 570 | -0.01 (-0.09 to 0.07) | 0.79 |
| | Ordinal | Child | 6,033 | 1.02 (0.97 to 1.07) | 0.48 |
| Cabbage intake | Ordinal | Mother (T1) | 3,451 | 1.04 (0.97 to 1.11) | 0.24 |
| | | Mother (T2) | 3,007 | 1.02 (0.95 to 1.09) | 0.63 |
| | | Father | 818 | 1.03 (0.90 to 1.18) | 0.65 |
| | | Child (T1) | 6,058 | 1.02 (0.97 to 1.07) | 0.39 |
| | | Child (T2) | 5,973 | 1.00 (0.95 to 1.05) | 0.90 |
| Mobile phone usage | Ordinal | Mother | 1,235 | 0.94 (0.85 to 1.05) | 0.26 |
| | | Father | 331 | 1.00 (0.83 to 1.22) | 0.98 |
| | | Child (weekday) | 4,208 | 1.02 (0.96 to 1.09) | 0.52 |
| | | Child (weekend day) | 4,444 | 1.03 (0.97 to 1.10) | 0.28 |
| Mother's age at time of questionnaire | Linear | Mother | 2,972 | -0.06 (-0.25 to 0.14) | 0.58 |
| | | Father | 685 | 0.19 (-0.22 to 0.61) | 0.36 |
| Back pain | Logistic | Mother (T1) | 3,425 | 0.99 (0.92 to 1.06) | 0.81 |
| | | Mother (T2) | 3,358 | 0.98 (0.92 to 1.05) | 0.60 |
| | | Father | 772 | 1.00 (0.86 to 1.16) | 0.96 |
| ***Smoking heaviness PRS – ever smokers*** | | | | | |
| Age at first pregnancy | Linear | Mother | 3,471 | -0.07 (-0.21 to 0.07) | 0.33 |
| Townsend deprivation index | Ordinal | Mother (T1) | 2,091 | 0.94 (0.87 to 1.02) | 0.13 |
| | | Mother (T2) | 2,864 | 0.95 (0.89 to 1.01) | 0.12 |
| Attends a place of worship | Ordinal | Mother | 3,062 | 0.89 (0.83 to 0.96) | 0.003 |
| | | Father | 629 | 1.00 (0.85 to 1.17) | 0.97 |
| Oat cereal intake | Ordinal | Mother (T1) | 3,129 | 0.98 (0.92 to 1.04) | 0.52 |
| | | Mother (T2) | 2,559 | 1.01 (0.93 to 1.10) | 0.77 |

| | | | | | |
|---|---|---|---|---|---|
| | | Father | 476 | 1.04 (0.86 to 1.25) | 0.70 |
| Wholegrain/bran cereal intake | Ordinal | Mother (T1) | 3,129 | 0.98 (0.92 to 1.06) | 0.06 |
| | | Mother (T2) | 2,559 | 0.98 (0.87 to1.06) | 0.65 |
| | | Father | 476 | 0.93 (0.79 to 1.10) | 0.39 |
| Other cereal intake | Ordinal | Mother (T1) | 3,129 | 1.03 (0.96 to 1.10) | 0.39 |
| | | Mother (T2) | 2,559 | 1.04 (0.97 to 1.12) | 0.24 |
| | | Father | 472 | 0.94 (0.80 to 1.11) | 0.47 |
| Time spent watching television (weekday) | Linear | Mother | 1,956 | 0.02 (-0.02 to 0.07) | 0.35 |
| | | Father | 435 | -0.07 (-0.16 to 0.03) | 0.52 |
| Time spent watching television (weekend day) | Linear | Mother | 1,824 | 0.01 (-0.03 to 0.06) | 0.54 |
| | | Father | 411 | -0.03 (-0.12 to 0.06) | 0.52 |
| Cabbage intake | Ordinal | Mother (T1) | 3,129 | 1.04 (0.97 to 1.11) | 0.31 |
| | | Mother (T2) | 2,559 | 1.02 (0.94 to 1.10) | 0.70 |
| | | Father | 639 | 1.07 (0.93 to 1.25) | 0.34 |
| Mobile phone usage | Ordinal | Mother | 1,027 | 0.95 (0.84 to 1.07) | 0.39 |
| | | Father | 275 | 0.96 (0.76 to 1.21) | 0.72 |
| Mother's age at time of questionnaire | Linear | Mother | 2,613 | -0.07 (-0.28 to 0.15) | 0.54 |
| | | Father | 528 | 0.312 (-0.16 to 0.79) | 0.19 |
| Back pain | Logistic | Mother (T1) | 3,069 | 1.01 (0.94 to 1.10) | 0.71 |
| | | Mother (T2) | 2,945 | 0.93 (0.87 to 1.01) | 0.07 |
| | | Father | 574 | 0.97 (0.83 to 1.15) | 0.76 |
| ***Smoking initiation PRS*** | | | | | |
| Age at first pregnancy | Linear | Mother | 7,199 | -0.15 (-0.24 to -0.06) | 0.001 |
| Townsend deprivation index | Ordinal | Mother (T1) | 4,376 | 1.04 (0.99 to 1.10) | 0.13 |
| | | Mother (T2) | 6,174 | 1.02 (0.98 to 1.07) | 0.33 |
| Attends a place of worship | Ordinal | Mother | 6,613 | 0.92 (0.88 to 0.96) | 0.0004 |
| | | Father | 1,442 | 0.94 (0.85 to 1.04) | 0.20 |
| | | Child | 5,110 | 0.95 (0.90 to 1.00) | 0.05 |
| Oat cereal intake | Ordinal | Mother (T1) | 6,712 | 1.00 (0.96 to 1.04) | 0.94 |
| | | Mother (T2) | 5,662 | 0.94 (0.89 to 0.99) | 0.01 |
| | | Father | 1,155 | 0.90 (0.80 to 1.01) | 0.07 |

| | | | | | |
|---|---|---|---|---|---|
| | | Child | 5,671 | 0.98 (0.93 to 1.03) | 0.44 |
| Wholegrain/bran cereal intake | Ordinal | Mother (T1) | 6,712 | 0.96 (0.92 to 1.00) | 0.08 |
| | | Mother (T2) | 5,662 | 0.96 (0.92 to 1.01) | 0.09 |
| | | Father | 1,156 | 0.98 (0.88 to 1.08) | 0.66 |
| | | Child | 5,776 | 1.00 (0.95 to 1.05) | 0.96 |
| Other cereal intake | Ordinal | Mother (T1) | 6,712 | 1.03 (0.99 to 1.08) | 0.15 |
| | | Mother (T2) | 5,662 | 0.98 (0.94 to 1.03) | 0.45 |
| | | Father | 1,149 | 0.95 (0.86 to 1.06) | 0.37 |
| | | Child | 5,901 | 1.01 (0.96 to 1.06) | 0.78 |
| Risk taking (higher = avoids risks more) | Ordinal | Child (T1) | 6,023 | 0.93 (0.88 to 0.97) | 0.001 |
| | | Child (T2) | 5,991 | 0.95 (0.91 to 1.00) | 0.04 |
| Time spent watching television (weekday) | Linear | Mother | 4,426 | 0.02 (-0.01 to 0.04) | 0.27 |
| | | Father | 1,066 | -0.03 (-0.08 to 0.03) | 0.39 |
| | Ordinal | Child | 6,072 | 1.01 (0.96 to 1.06) | 0.69 |
| Time spent watching television (weekend day) | Linear | Mother | 4,129 | 0.02 (-0.008 to 0.05) | 0.15 |
| | | Father | 1,019 | -0.02 (-0.08 to 0.04) | 0.51 |
| | Ordinal | Child | 6,033 | 1.00 (0.96 to 1.05) | 0.88 |
| Cabbage intake | Ordinal | Mother (T1) | 6,712 | 1.01 (0.96 to 1.06) | 0.64 |
| | | Mother (T2) | 5,661 | 0.99 (0.94 to 1.04) | 0.68 |
| | | Father | 1,463 | 1.01 (0.92 to 1.11) | 0.86 |
| | | Child (T1) | 6,058 | 1.05 (1.01 to 1.11) | 0.02 |
| | | Child (T2) | 5,973 | 0.99 (0.95 to 1.04) | 0.74 |
| Mobile phone usage | Ordinal | Mother | 2,306 | 1.08 (1.00 to 1.16) | 0.05 |
| | | Father | 630 | 1.01 (0.87 to 1.17) | 0.88 |
| | | Child (weekday) | 4,208 | 1.01 (0.95 to 1.08) | 0.73 |
| | | Child (weekend day) | 4,444 | 1.04 (0.98 to 1.10) | 0.16 |
| Mother’s age at time of questionnaire | Linear | Mother | 5,688 | -0.16 (-0.30 to -0.02) | 0.03 |
| | | Father | 1,217 | -0.35 (-0.67 to -0.03) | 0.03 |
| Back pain | Logistic | Mother (T1) | 6,643 | 1.02 (0.97 to 1.07) | 0.50 |
| | | Mother (T2) | 6,452 | 1.00 (0.95 to 1.05) | 1.00 |
| | | Father | 1,388 | 1.04 (0.93 to 1.16) | 0.46 |

For linear regression models the effect estimate is beta and for logistic and ordinal regressions this is the odds ratio. OR=odds ratio, CI=confidence interval, PRS=polygenic risk score. T1 = time point 1, T2 = time point 2

**Supplementary Fig S1. Associations between polygenic risk scores for lifetime smoking score (blue), smoking heaviness (red, from darkest to lightest is never, former and current) and smoking initiation (green) and plausible phenotypes (linear models)**

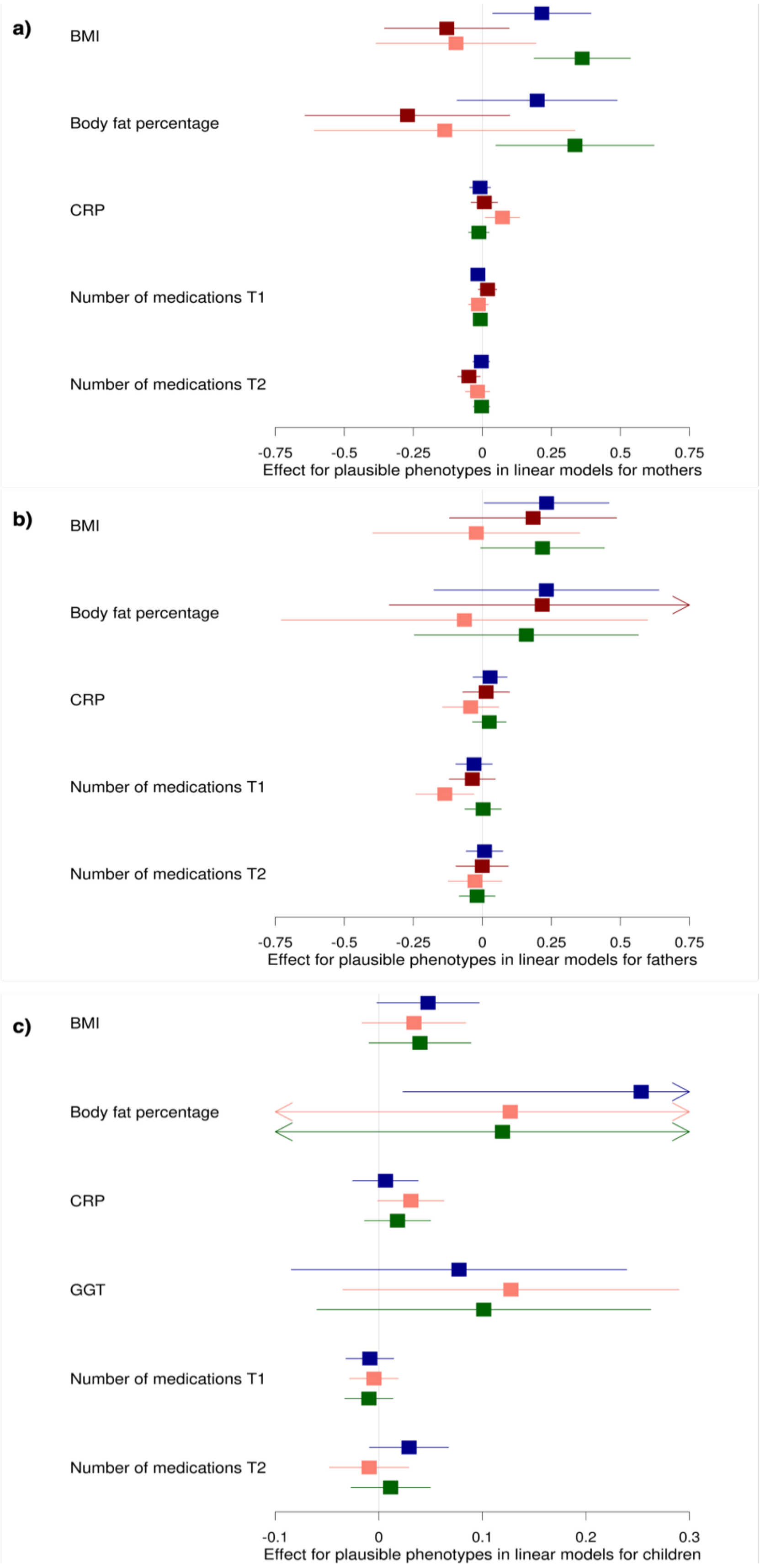

Associations between polygenic risk scores for lifetime smoking index (blue), smoking heaviness (red, from darkest to lightest is never and ever, except for children where there is a single result) and smoking initiation (green) and plausible phenotypes from linear models. Results are presented for mothers (a), fathers/partners (b) and children (c). The effect estimate is beta for linear regressions. BMI=body mass index, CRP=C-reactive protein, GGT=Gamma Glutamyl Transferase, GP=general practitioner.

**Supplementary Fig S2. Associations between polygenic risk scores for lifetime smoking score (blue), smoking heaviness (red, from darkest to lightest is never, former and current) and smoking initiation (green) and plausible phenotypes (logistic models)**

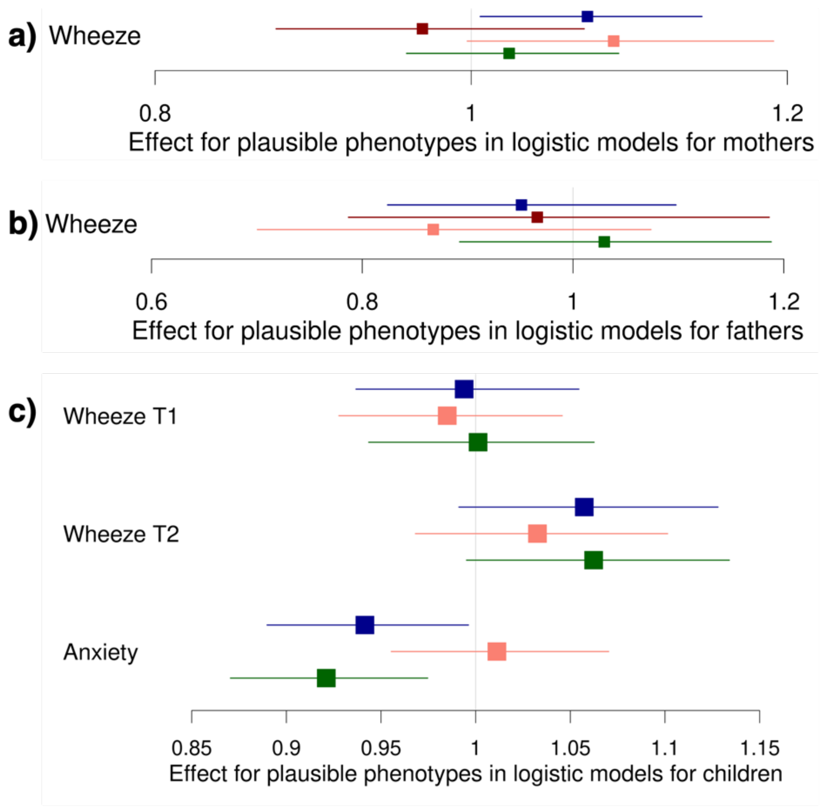


Associations between polygenic risk scores for lifetime smoking index (blue), smoking heaviness (red, from darkest to lightest is never and ever, except for children where there is a single result) and smoking initiatin (green) and plausible from logistic models. Results are presented for mothers (a), fathers/partners (b) and children (c). The effect estimate is odds ratios for logistic regressions.

**Supplementary Fig S3. Associations between polygenic risk scores for lifetime smoking score (blue), smoking heaviness (red, from darkest to lightest is never, former and current) and smoking initiation (green) and plausible phenotypes (ordinal models)**

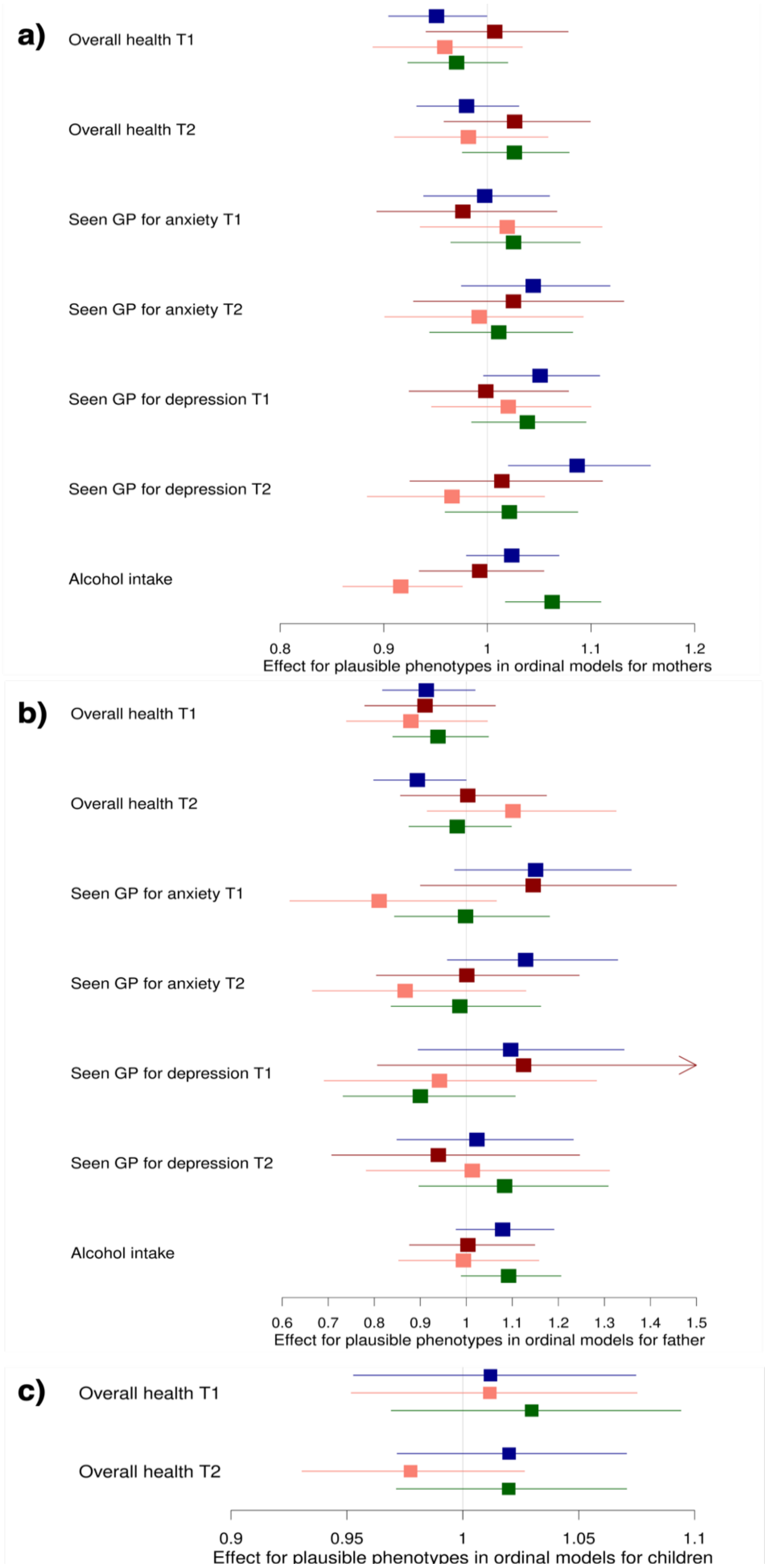

Associations between polygenic risk scores for lifetime smoking index (blue), smoking heaviness (red, from darkest to lightest is never and ever, except for children where there is a single result) and smoking initiation (green) and plausible from ordinal models. Results are presented for mothers (a), fathers/partners (b) and children (c). The effect estimate is odds ratios for ordinal regressions.

**Supplementary Fig S4. Associations between polygenic risk scores for lifetime smoking score (blue), smoking heaviness (red, from darkest to lightest is never, former and current) and smoking initiation (green) and implausible phenotypes (linear models)**

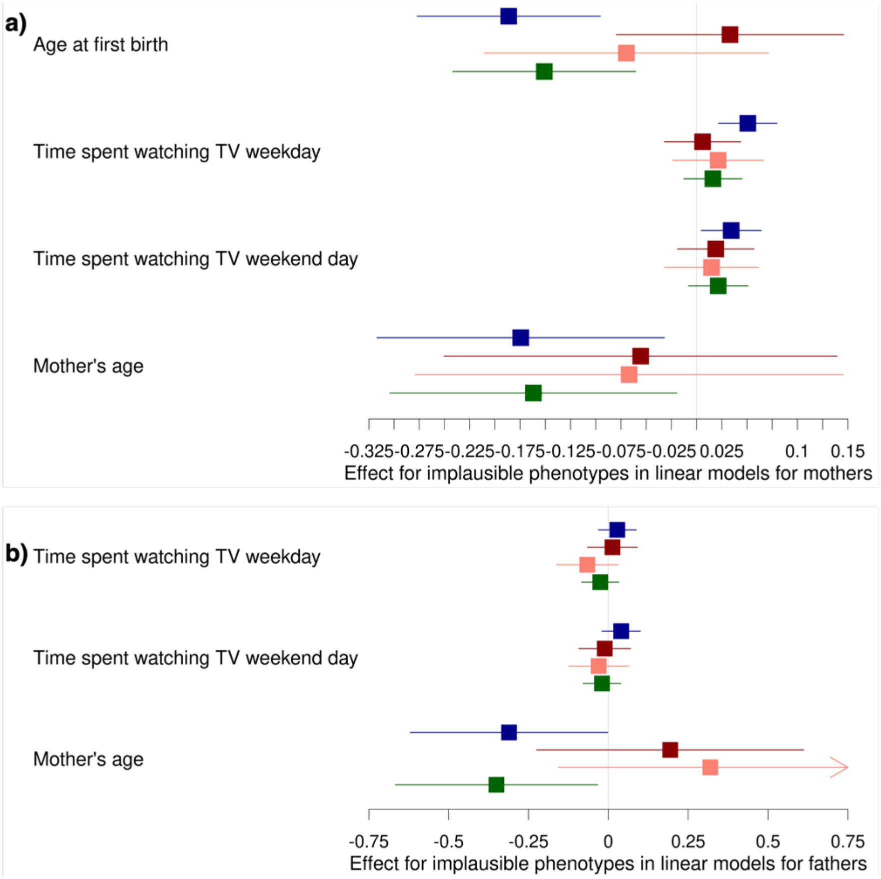


Associations between polygenic risk scores for lifetime smoking index (blue), smoking heaviness (red, from darkest to lightest is never and ever, except for children where there is a single result) and smoking initiation (green) and implausible phenotypes from linear models. Results are presented for mothers (a), fathers/partners (b) and children (c). The effect estimate is beta for linear regressions. BMI=body mass index, CRP=C-reactive protein, GGT=Gamma Glutamyl Transferase, GP=general practitioner.

**Supplementary Fig S5. Associations between polygenic risk scores for lifetime smoking score (blue), smoking heaviness (red, from darkest to lightest is never, former and current) and smoking initiation (green) and implausible phenotypes (logistic models)**

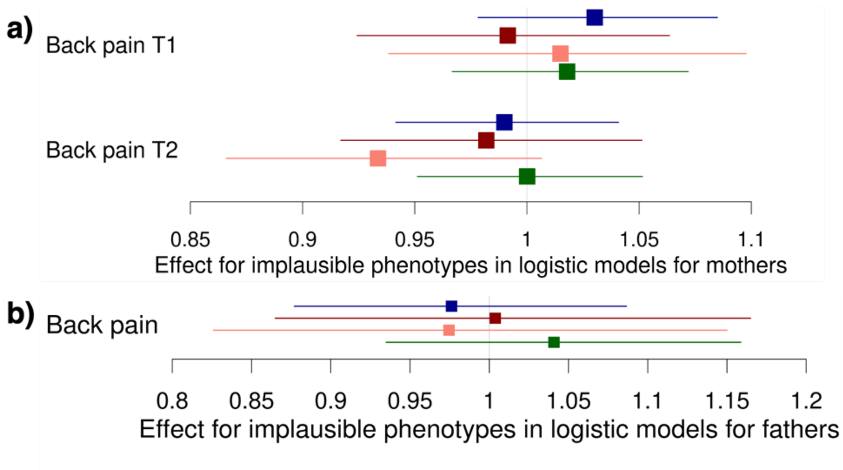


Associations between polygenic risk scores for lifetime smoking index (blue), smoking heaviness (red, from darkest to lightest is never and ever, except for children where there is a single result) and smoking initiation (green) and plausible and implausible phenotypes from logistic models. Results are presented for mothers (a), fathers/partners (b) and children (c). The effect estimate is odds ratios for logistic regressions.

**Supplementary Fig S6. Associations between polygenic risk scores for lifetime smoking score (blue), smoking heaviness (red, from darkest to lightest is never, former and current) and smoking initiation (green) and implausible phenotypes (ordinal models)**

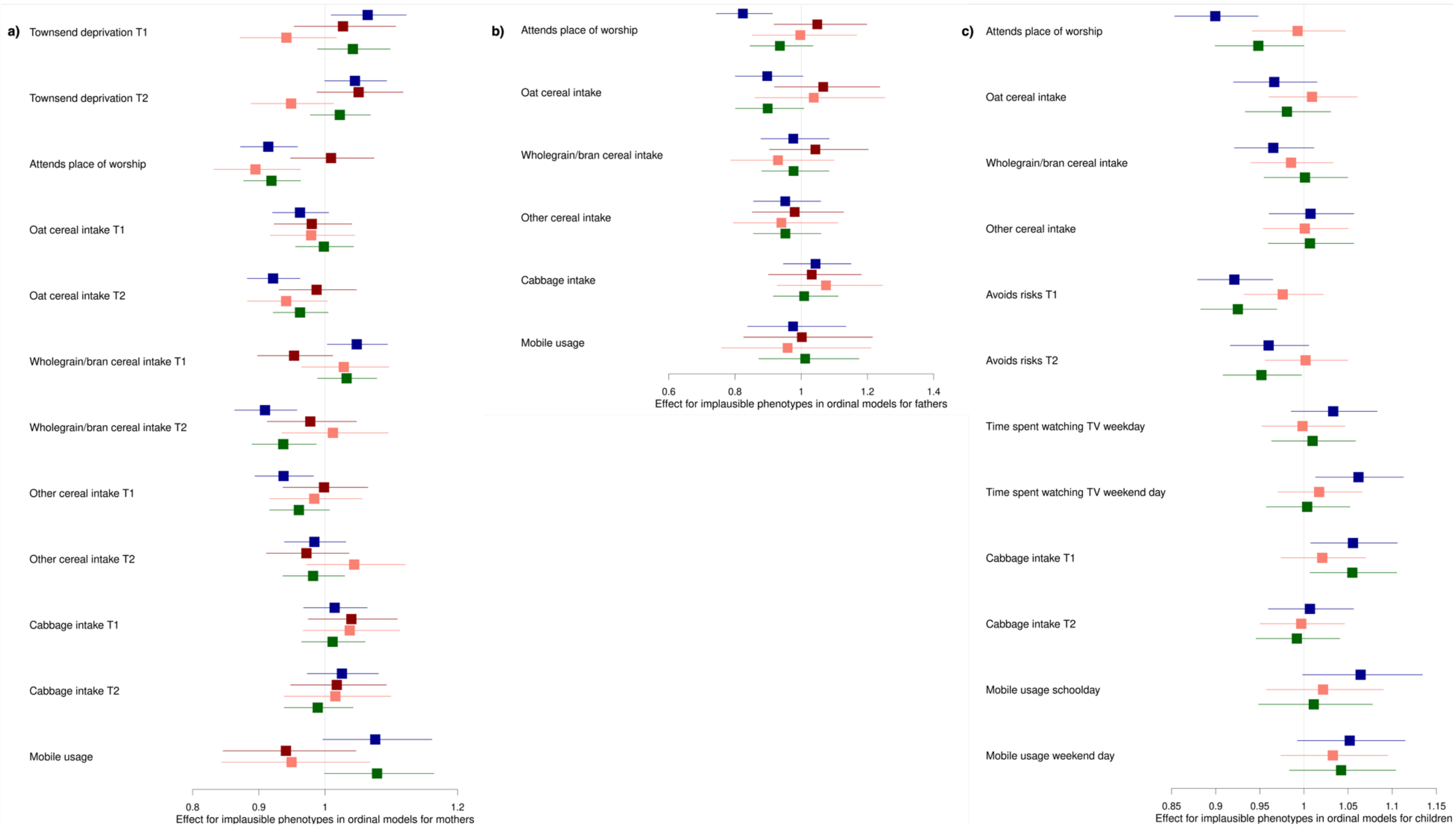


Associations between polygenic risk scores for lifetime smoking index (blue), smoking heaviness (red, from darkest to lightest is never and ever, except for children where there is a single result) and smoking initiation (green) and implausible phenotypes from ordinal models. Results are presented for mothers (a), fathers/partners (b) and children (c). The effect estimate is odds ratios for ordinal regressions.
